# Genetic studies of gestational diabetes mellitus in 21,813 Chinese women

**DOI:** 10.1101/2023.11.23.23298977

**Authors:** Huanhuan Zhu, Han Xiao, Linxuan Li, Meng Yang, Mingzhi Cai, Jieqiong Zhou, Jingyu Zeng, Yan Zhou, Xianmei Lan, Jiuying Liu, Xiaobo Qian, Yuanyuan Zhong, Panhong Liu, Xinyi Zhang, Ying Lin, Zhongqiang Cao, Hong Mei, Xiaonan Cai, Liqin Hu, Rui Zhou, Xun Xu, Huanming Yang, Jian Wang, Xin Jin, Aifen Zhou

**Affiliations:** BGI-Shenzhen, Shenzhen, China; Institute of Maternal and Child Health, Wuhan Children’s Hospital (Wuhan Maternal and Child Health care Hospital), Tongji Medical College, Huazhong University of Science and Technology, Wuhan, China; College of Life Sciences, University of Chinese Academy of Sciences, Beijing 100049, China; Department of Obstetrics, Wuhan Children’s Hospital (Wuhan Maternal and Child Health care Hospital), Tongji Medical College, Huazhong University of Science and Technology, Wuhan, China; College of Life Sciences, Northwest A&F University, Yangling, Shaanxi, China; BGI-Wuhan Clinical Laboratories, BGI-Shenzhen, Wuhan, China; Guangdong Provincial Key Laboratory of Genome Read and Write, Shenzhen, China; Guangdong Provincial Academician Workstation of BGI Synthetic Genomics, Shenzhen, China; James D. Watson Institute of Genome Sciences, Hangzhou, China; The Cancer Hospital of the University of Chinese Academy of Sciences (Zhejiang Cancer Hospital), Institute of Basic Medicine and Cancer (IBMC), Chinese Academy of Sciences, 16 Hangzhou, China; BGI, BGI-Shenzhen, Shenzhen, China; School of Medicine, South China University of Technology, Guangzhou, China

## Abstract

Gestational diabetes mellitus (GDM) is a common complication of pregnancy with adverse effects on both mothers and fetuses. The relationship between GDM and clinical information has been largely investigated; however, the role of laboratory features and genetic components in GDM are relatively understudied, especially in the Chinese population. In this study, we recruited 21,813 pregnant Chinese women and investigated the risk factors for GDM from both environmental and genetic aspects. In addition to the serum fasting glucose and urine glucose measured in the first trimester, our study revealed the close relations of γ-glutamyl transferase, prealbumin, and uric acid to GDM. The GWAS and subsequent conditional & joint association analysis identified four genome-wide significant SNPs, rs35261542, rs4712530, rs3781637, and rs12225378, mapped on *CDKAL1* and *MTNR1B*. The allele frequencies of the latter three SNPs showed substantial differences in European and East Asian populations. The ancient DNA analysis demonstrated that mutation of rs3781637 was originally appeared in an ancient Chinese in the Holocene period. This result might provide useful reference for explaining the distinct genetic background of GDM in different populations. Considering both clinical measurements and genetic components, we constructed an early prediction model for GDM by using only a dozen indicators and achieved acceptable prediction performance. We believe that our study will certainly become an important reference for the genetic study of GDM and will have important implications for elucidating the genetic mechanisms of GDM.

## Background

Gestational diabetes mellitus (GDM) is one of the common complications of pregnancy in women with normal glucose metabolism before pregnancy while developing hyperglycemic symptoms and being diagnosed with diabetes during pregnancy. Due to the lack of uniform screening modalities and diagnostic criteria, the prevalence of GDM varies widely worldwide, ranging from 1% to 30%^1^. In China, the GDM prevalence is as high as 14.8%, and with the improvement of living standards, this rate is growing every year^2^. GDM can cause a variety of adverse outcomes to both maternal and offspring health, such as preeclampsia, miscarriage, premature delivery, and fetal malformation^3–5^. In 2019, the International Diabetes Federation reported that about 1/6 of live births worldwide (20 million) are affected by hyperglycemia^6^, which imposes a heavy burden on both families and society.

Numerous studies have shown that older maternal age, overweight or obesity, excessive weight gain during pregnancy, polycystic ovary syndrome, multiple pregnancies, excessive amniotic fluid, previous history of GDM, and family history of GDM are important risk factors for GDM^7–10^. The incidence and course of GDM are influenced by both environmental and genetic risk factors. In the last twenty years, genome-wide association studies (GWAS) have been used to identify genomic variants that are statistically associated with the disease of interest^11^. As of March 2023, the GWAS Catalog have compiled six GWAS analysis for GDM in different populations. The first one was published in 2012, which recruited 1,710 Korean pregnant women and identified two genome-wide significant loci (*MTNR1B* and *CDKAL1*) ^12^. The other studies were issued in recent years, including one Chinses study ^13^, two European studies^14,15^, a mixed of multiple populations ^16^, and European population with FinnGen and UK Biobank participants^17^. Neither the Chinese study or the two European studies uncovered genome-wide significant variants. The mixed population study, including 8,185 Asian subjects, replicated *MTNR1B* and *CDKAL1*, and for the first time found *TCF7L2*, *CDKN2B-AS1*, and *HKDC1*. The meta-analysis of UK Biobank and FinnGen data, including 157 UKB cases and 7,238 Finnish cases, replicated *MTNR1B* and *TCF7L2*, and discovered *HLA-DQB1*.

In recent years, Non-Invasive Prenatal Testing (NIPT) has effectively reduced the birth rate of children with common fetal autosomal trisomies^18^. NIPT is a method for sequencing cell-free DNA fragments (including fetal cell-free DNA) in maternal peripheral plasma by obtaining maternal venous blood and using next-generation DNA sequencing technology. With the increasing popularity of NIPT technology, the prenatal screening of fetuses for chromosomal disorders has also generated a large amount of maternal genotype data, with which a series of genomic and population genetic analyses have been carried out^19^, a previous work of our group. In that work, we successfully performed genome-wide association analysis, viral infections, and population history analysis based on NIPT sequencing data of 141,431 pregnant Chinese women.

In this study, we explored the genetic background of GDM based on NIPT data from 21,813 Chinses women. We defined GDM case/control and low/high risk of GDM as the principal traits, and serum glucose levels in oral glucose tolerance test and the derived measurements as the secondary traits. First, we examined the statistical inference for clinical information and the principal traits, aiming to find the risk factors and adverse outcomes of GDM. The results were consistent with previous findings. Second, we performed GWAS on both principal and secondary traits. For the principal traits, we replicated *CDKAL1* and *MTNR1B* with independent variants rs35261542 and rs3781637, respectively. The ancient DNA analysis found that the original mutation of rs3781637 was appeared in an ancient Chinese man and its allele frequency is higher in Asian than European. This observation might provide useful hint for exploring the different genetic background of GDM across populations. Third, we further carried out gene-based and protein-protein interaction (PPI) analysis based on the GWAS results. The PPI enrichment analysis identified several groups of significant pathways, including glucose metabolism, hormone levels, and postnatal outcomes. Finally, we constructed predictive models for the principal GDM traits by considering both environmental and genetic risk factors. The averaged AUC in GDM prediction model reached 76%. Our work is the largest genetic study of GDM in the Asian population to date and hopefully will provide an important reference for revealing the disease etiology of GDM and the different genetic mechanisms in Asian and European population.

## Results

### Sample participants

All the subjects in this study were recruited from Wuhan Children’s Hospital, located in Wuhan city, Hubei province, Central China. In detail, 39,194 pregnant women took NIPT tests and produced the sequencing data. After quality control, 38,668 subjects were filtered in for genotype imputation. On average, the imputation accuracy was 81.37% and the data was eligible for genome-wide association analysis. More details were provided in our companion work [cite].

### Screening for the principal GDM traits

Based on the hospital’s electronic information system, a total of 21,813 subjects underwent the OGTT tests and 2,502 were diagnosed as GDM cases, the disease prevalence being 11.47% (Figure 1a). The numbers of GDM cases with the criteria of fasting plasma glucose (≥5.1 mmol/L), 1-hour glucose (≥10.0 mmol/L), and 2-hour glucose (≥8.5 mmol/L) were 1,055, 1,127, and 1,404, respectively. The distributions of glucose levels in GDM-control and GDM-case were provided in Figure 1a. Unsurprisingly, patients in the case group had significantly higher glucose levels than the control group.

**Figure 1.**
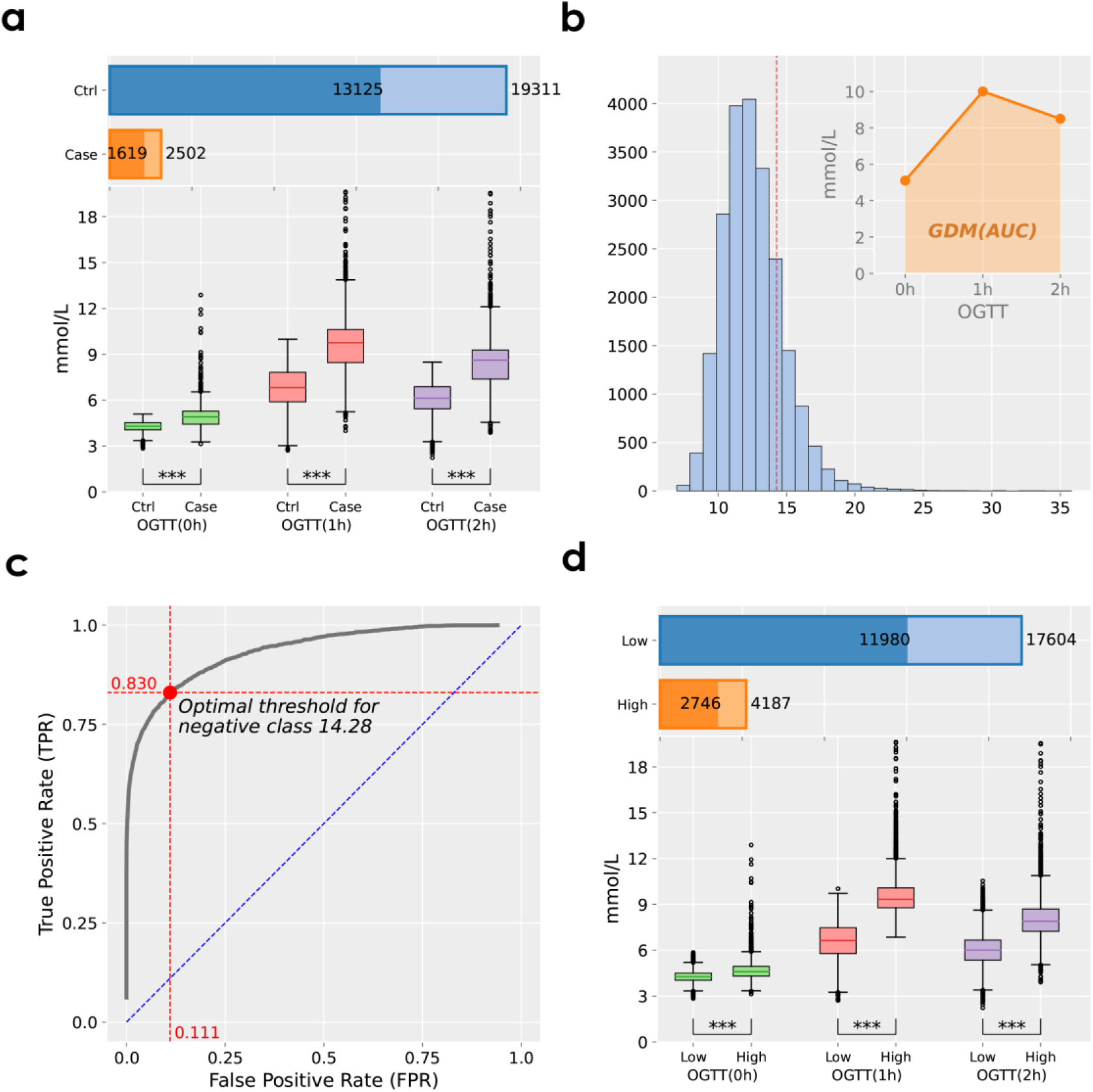
Screening for the principal GDM traits *Notes*: **a)** the numbers of GDM control and case in bar plots, bars in light color provide all available samples sizes and bars in dark color provide effective sample sizes in subsequent GWAS analysis; the boxplots of plasma glucose levels in the OGTT test for GDM control (Ctrl) and case, *** means p-value was less than 0.001. **b)** the distribution of GDM_AUC_ (mmol*h/L), which was calculated as the area under the three plasma glucose levels. **c)** the ROC curve showing how to choose the optimal threshold of GDM_AUC_ for dichotomizing high- and low-risk samples. **d)** the numbers of high- and low-risk of GDM in bar plots; the boxplots of glucose levels in the OGTT test for high- and low-risk samples.

The histogram of GDM_AUC_ was provided in Figure 1b. We then classified the individuals into GDM high-risk and low-risk by dichotomizing the GDM_AUC_ with a grid of thresholds ranging from 9.5 to 20 with an increment of 0.01. For each threshold, we calculated the false positive rate (FPR) and true positive rate (TPR). The generated ROC curve was provided in Figure 1c. By optimizing the Youden’s J statistic, it produced the optimal threshold for high-low risk classification in 14.28. This value was quite consistent with a previously suggested threshold of 14.2^20^. At the threshold of AUC >14.28, 4,187 samples were classified as GDM high-risk, while 17,604 samples were in low-risk, and the rate of GDM high-risk was 19.21% (Figure 1d). We also provided the distributions of OGTT glucose levels in high- and low-risk groups (Figure 1d), which had the same pattern as GDM-case and control groups.

### Associated environmental factors of the principal traits

The statistical inference of GDM_status_ and GDM_risk_ on clinical information are quite consistent (Table 1, Supplementary Table S1). In brief, advanced maternal age, obesity, and high blood pressure are inferred as significant risk factors for GDM. On the other hand, women with GDM tend to have shorter gestational lengths, deliver preterm infants with low birthweight or macrosomic infant, and suffer cesarean section. When the mother is diagnosed with GDM, there is no significant difference for being premature between female and male fetuses (OR = 0.90, p-value = 0.51 from chi-square test), while the male fetus is more likely to be macrosomic than the female fetus (OR = 1.88, p-value = 0.00028 from chi-square test).

**Table 1.** Regression analysis between clinical information and GDM_status_.

| Group | Characteristic | All samples | Control | Case | t/OR | p-value |
| --- | --- | --- | --- | --- | --- | --- |
|  |  | (N=21,813) | (N=19,311) | (N=2,502) |  |  |
| Risk factors | Age (years) | 29.58±4.03 | 29.37±3.94 | 31.20±4.32 | 21.593 | **2.45E-102** |
|  | Weight (kg) | 55.94±12.34 | 55.51±12.59 | 59.18±9.75 | 13.33 | **2.35E-40** |
|  | BMI (kg/m<sup>2</sup>) | 21.39±3.08 | 21.22±2.98 | 22.67±3.45 | 21.235 | **6.20E-99** |
|  | Obesity, n(%) | 2175(11.35%) | 1662(9.82%) | 513(22.79%) | 2.71 | **6.46E-74** |
|  | SBP (mmHg) | 109.53±11.55 | 109.06±11.37 | 113.02±12.24 | 15.382 | **4.49E-53** |
|  | DBP (mmHg) | 68.61±9.63 | 68.32±9.63 | 70.82±9.33 | 11.622 | **4.03E-31** |
|  | First-time pregnancy, n(%) | 7769(55.73%) | 7176(57.09%) | 593(43.25%) | 0.573 | **1.60E-22** |
| Postnatal outcomes | Gestational weeks (weeks) | 38.74±1.58 | 38.78±1.55 | 38.37±1.74 | -12.324 | **8.84E-35** |
|  | Premature birth, n(%) | 1206(5.53%) | 986(5.11%) | 220(8.80%) | 1.793 | **4.17E-14** |
|  | Birth weight (g) | 3286.43±473.71 | 3287.08±464.74 | 3281.42±537.96 | -0.561 | 5.75E-01 |
|  | Low birthweight, n(%) | 979(4.52%) | 821(4.28%) | 158(6.35%) | 1.517 | **3.59E-06** |
|  | Giant baby, n(%) | 1054(4.86%) | 889(4.63%) | 165(6.63%) | 1.462 | **1.60E-05** |
|  | C-section, n(%) | 11789(54.10%) | 10225(53.01%) | 1564(62.56%) | 1.481 | **2.31E-19** |
|  | Newborn gender, n(%) | 10418(48.07%) | 9218(48.05%) | 1200(48.25%) | 1.008 | 8.64E-01 |

The results of logistic regression between laboratory tests in the first trimester and the principal GDM traits were given in Figure 2a-b and Supplementary Table S2. Specifically, the first few significantly associated measurements are plasma glucose, γ -glutamyl transferase, prealbumin, urine glucose, uric acid, platelet count, and hemoglobin. Based on our prior knowledge, we expected the strong associations of serum and urinary glucose levels with GDM. Intriguingly, γ-glutamyl transferase, prealbumin, and uric acid were highly GDM-associated with p-values less than 1e-20. The distributions of these features also showed visible difference in case and control groups (Figure 2c-d). These results provided useful reference for clinical physicians and pregnant women that in addition to the serum and urinary glucose, considerable attentions should be paid to γ-glutamyl transferase, prealbumin, and uric acid.

**Figure 2.**
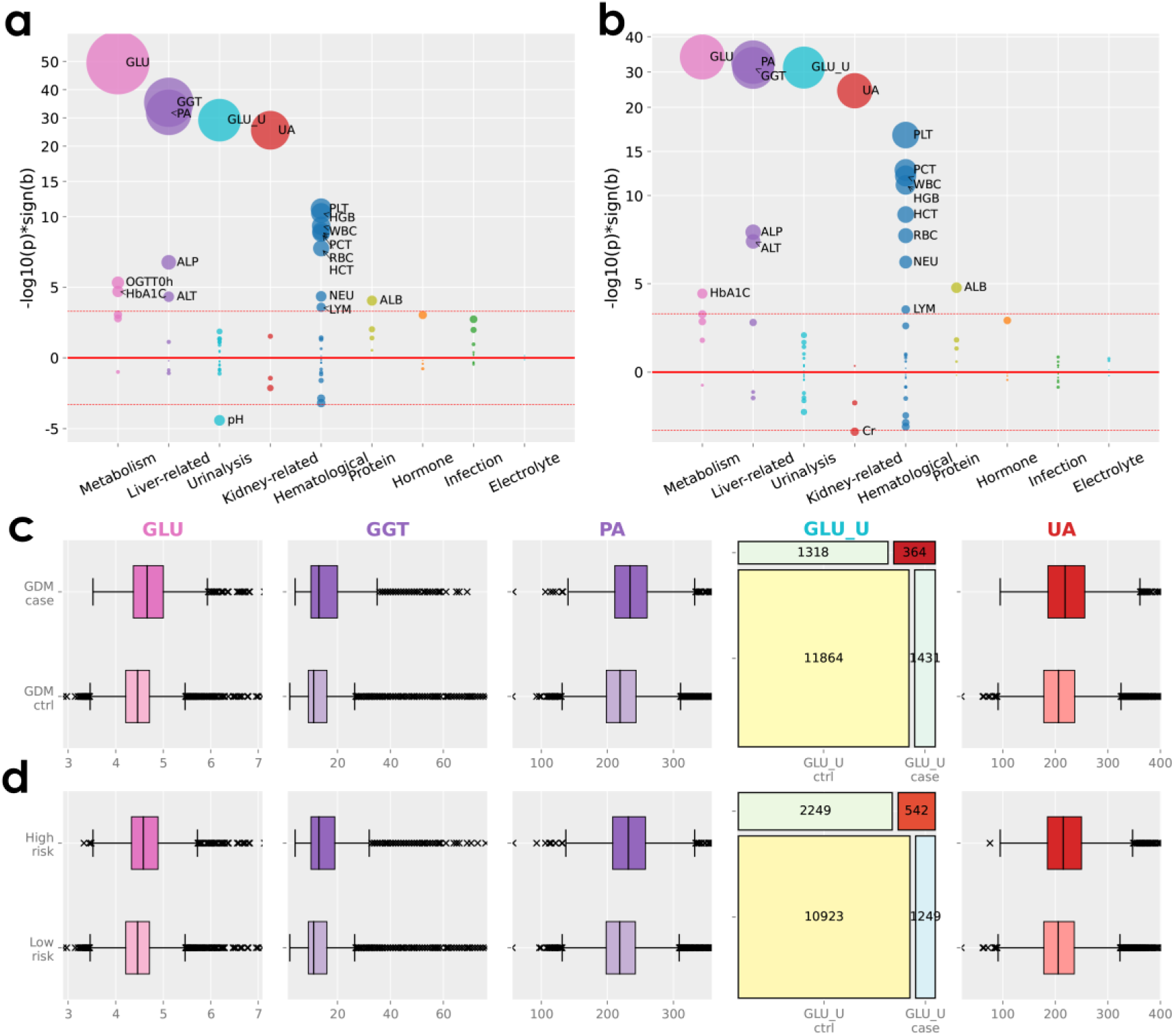
Regression analysis of the principal traits on laboratory features *Notes*: **a)** the bubble plot displays the results of regressing GDM_status_ on early pregnancy traits, the y-axis indicates -log10(p)*sign(b), where p is the p-value and b is the coefficient from the logistic regression model. The thick red line is y=0 and the light red lines are y = ±Log10(5e-4) = ±3.30. **b)** the bubble plot displays the results of regressing GDM_risk_ on early pregnancy traits. **c)** the boxplots and 2x2 contingency table of selected laboratory traits in GDM case and control groups. **d)** the boxplots and 2x2 contingency table of selected laboratory traits in high- and low-risk samples.

### GWAS analysis of pregnancy glycemic traits

We note that the number of samples who took OGTT tests and also had NIPT genotype data was 14,744, among which 1,619 were diagnosed as GDM cases (Figure 1a). Similarly, the effective sample size for GDM_risk_ is 14,726, among which 2,746 were in high GDM risk (Figure 1d). The GWAS analysis identified two genome-wide associated loci for GDM_status_ and GDM_risk_: *CDKAL1* and *MTNR1B* (Figure 3, Tables 2-3). *CDKAL1* (Cyclin-Dependent Kinase 5 regulatory subunit associated protein 1-like 1) has been reported to be associated with proinsulin conversion defect and glucose-stimulated insulin response defect ^21,22^. *MTNR1B* (Melatonin Receptor 1B) encodes one of the two high-affinity forms of a receptor for the hormone melatonin, which acts in glucose homeostasis^23^.

**Figure 3.**
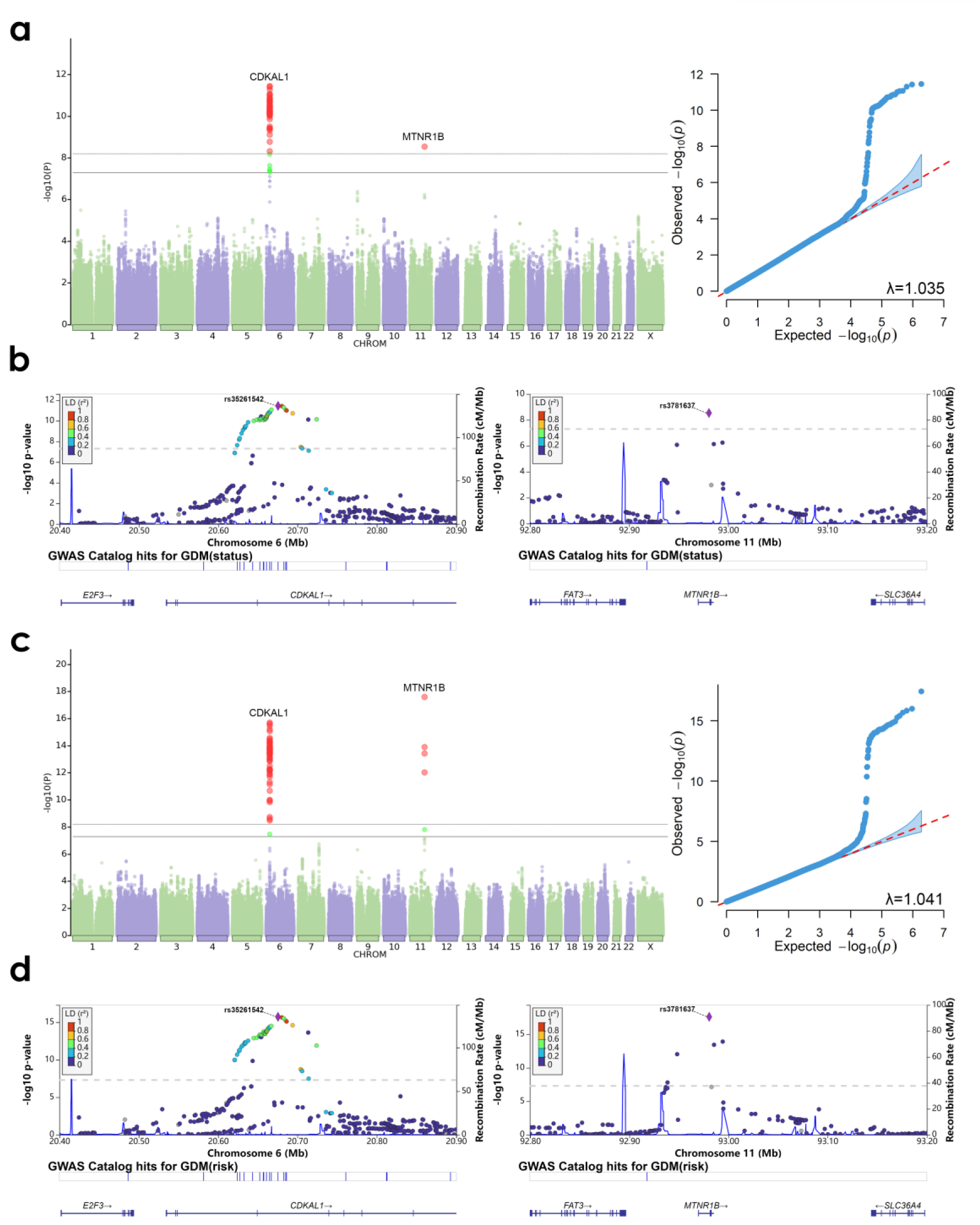
The GWAS results of the principal GDM traits *Notes*: **a)** Manhattan plot and QQ-plot with genomic inflation factor λ for GDM_status_, SNPs that pass the genome-wide and study-wide significance thresholds (5e-8 and 6.25e-9) were colored in green and red, respectively. **b)** LocusZoom plots of *CDKAL1* and *MTNR1B* association with GDMstatus. **c)** Manhattan plot and QQ-plot for GDM_risk_. **d)** LocusZoom plots of *CDKAL1* and *MTNR1B* association with GDMrisk.

**Table 2.**
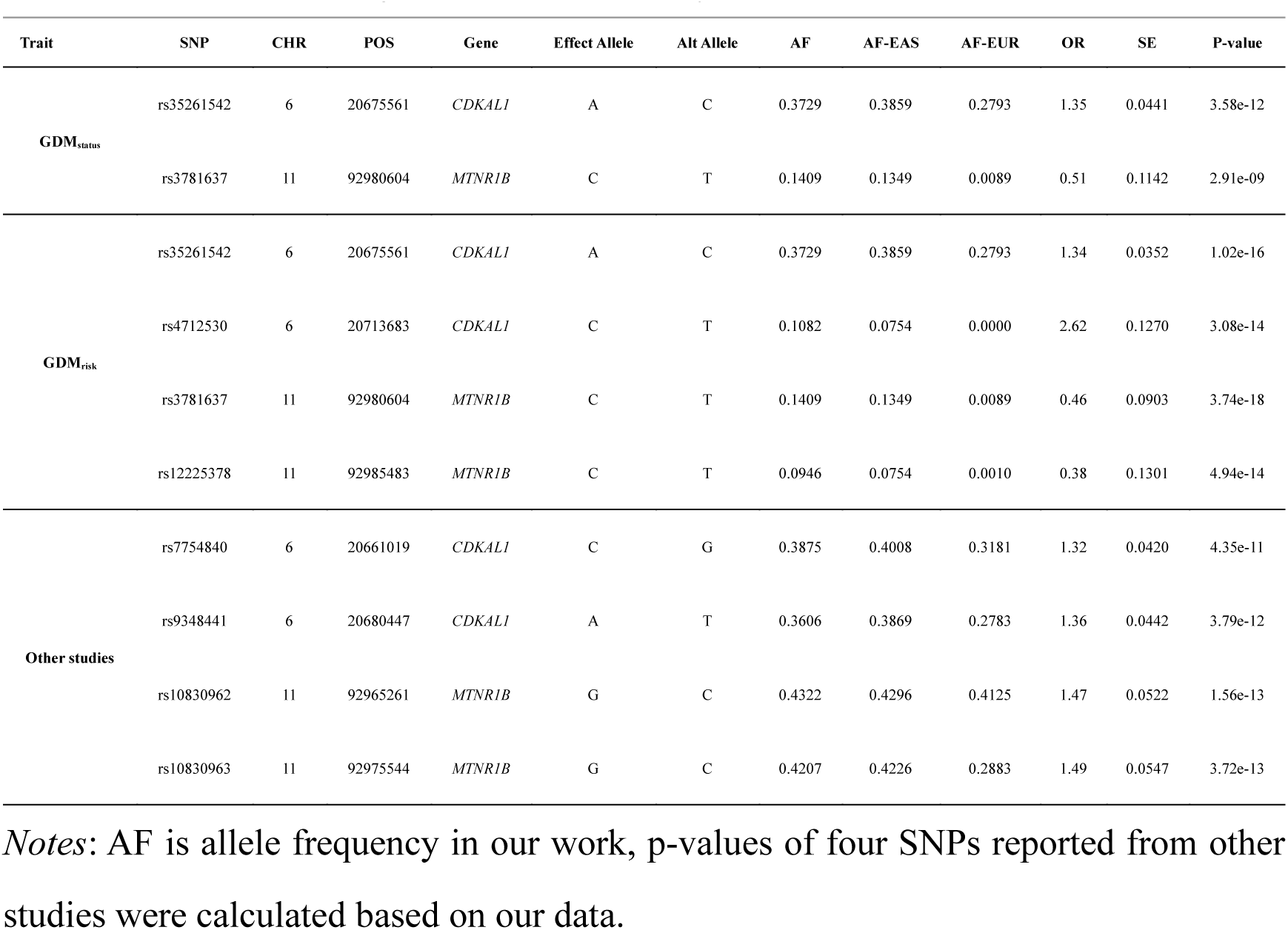
Conditional and joint association analysis results.

To identify independent signals, we further performed conditional and joint (COJO) association analysis in *CDKAL1* (chr6:20534457-21232404, GRCh38) and *MTNR1B* (chr11:92969651-92985066), respectively. This was implemented in GCTA with argument *--cojo-slct*. For GDM_status_, the COJO analysis outputted SNPs rs35261542 (chr6:20675561) and rs3781637 (chr11:92980604) (Table 2). For GDM_risk_, it additionally outputted rs4712530 (chr6:20713683) and rs12225378 (chr11:92985483) (Table 2). As mentioned in the Introduction section, three GWAS studies reported significant variants mapped on *CDKAL1* and *MTNR1B*, including rs7754840 (chr6:20661019), rs9348441 (chr6:20680447), rs10830962 (chr11:92965261), and rs10830963 (chr11:92975544). In this work, these SNPs were also genome-wide significantly associated with the principal traits (Table 2), even though SNPs rs10830962 and rs10830963 were filtered out from the GWAS summary statistics as they failed to pass Hardy-Weinberg equilibrium.

We provided the allele information for the aforementioned eight SNPs, including genomic coordinates, mapped gene, effect and alternate allele, allele frequencies in this work, in EAS, and EUR, and summary statistics (Table 2). We noticed that the allele frequencies of SNPs rs4712530, rs3781637, and rs12225378 have substantial differences between EAS and EUR, which might imply the different genetic background of GDM in different populations. The Geography of Genetic Variants (GGV, http://popgen.uchicago.edu/ggv/) also verified this observation (Figure 4, Supplementary Figure S1). To see if there are variants originally mutated from only EAS or EUR, we further performed ancient DNA analysis for the eight SNPs based on the Allen Ancient DNA Resource (AADR, https://reich.hms.harvard.edu/allen-ancient-dna-resource-aadr-downloadable-genotypes-present-day-and-ancient-dna-data). Four SNPs (rs3781637, rs7754840, rs10830962, rs10830963) were found in the AADR dataset. In particular, the mutation of SNP rs3781637 was initially appeared in an ancient Chinese in the Holocene period^24^ (Figure 5). This SNP was previously identified to be associated with T2D^25^ and tardive dyskinesia^26^ in Han Chinese population. Several evolutionary studies also investigated the natural selection at *MTNR1B*^27–29^. The mutations of other three SNPs did not display EAS or EUR disposition in origin (Supplementary Figure S2). For these eight SNPs, we also provided their allele frequency distributions in different Chinese provinces and regions (Supplementary Figure S3) calculated based on the CMDB database^30^.

**Figure 4.**
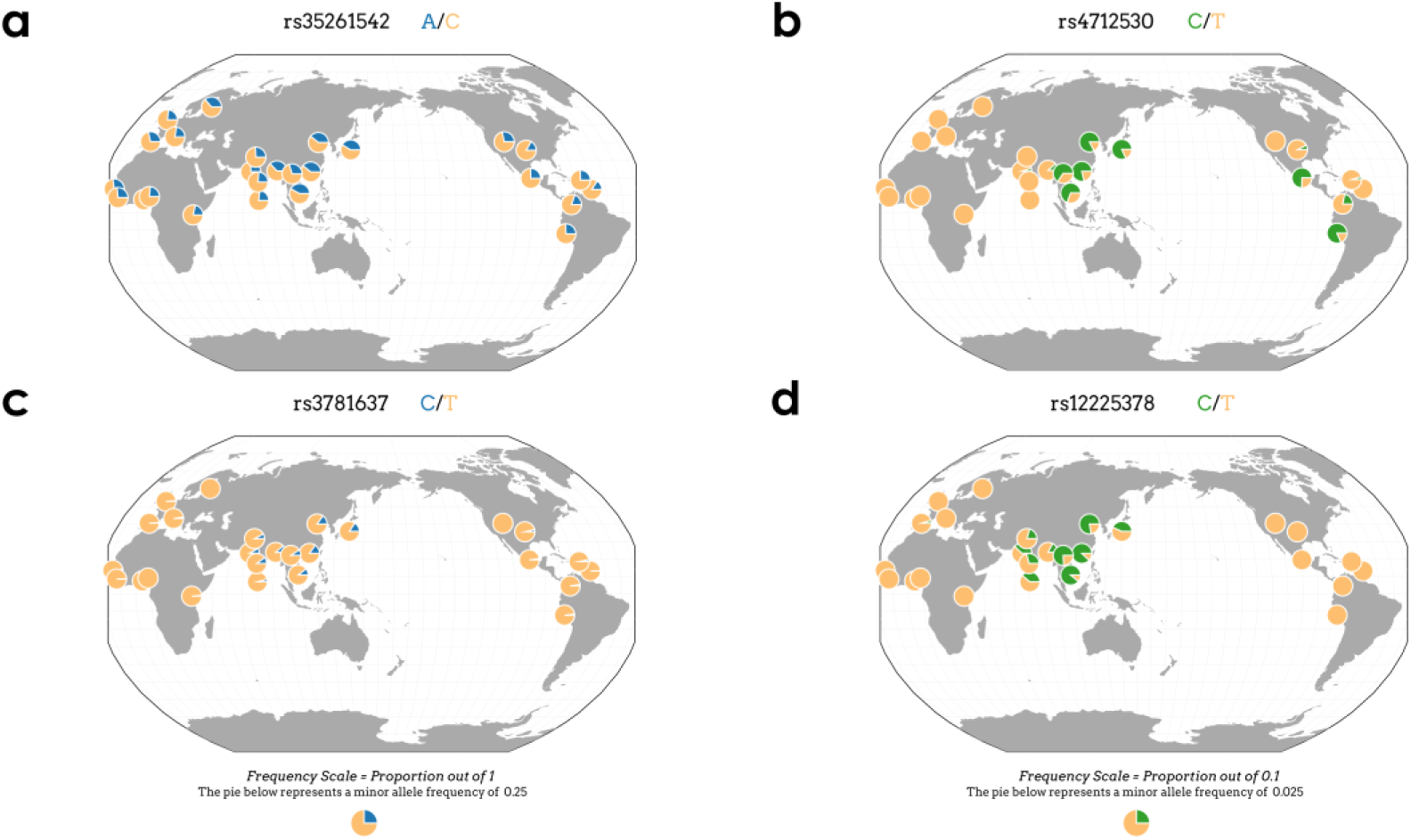
The global geographic distributions of identified genetic variants *Notes*: **a-d)** the geographic allele frequency distributions of rs35261542, rs4712530, rs3781637, and rs12225378.

**Figure 5.**
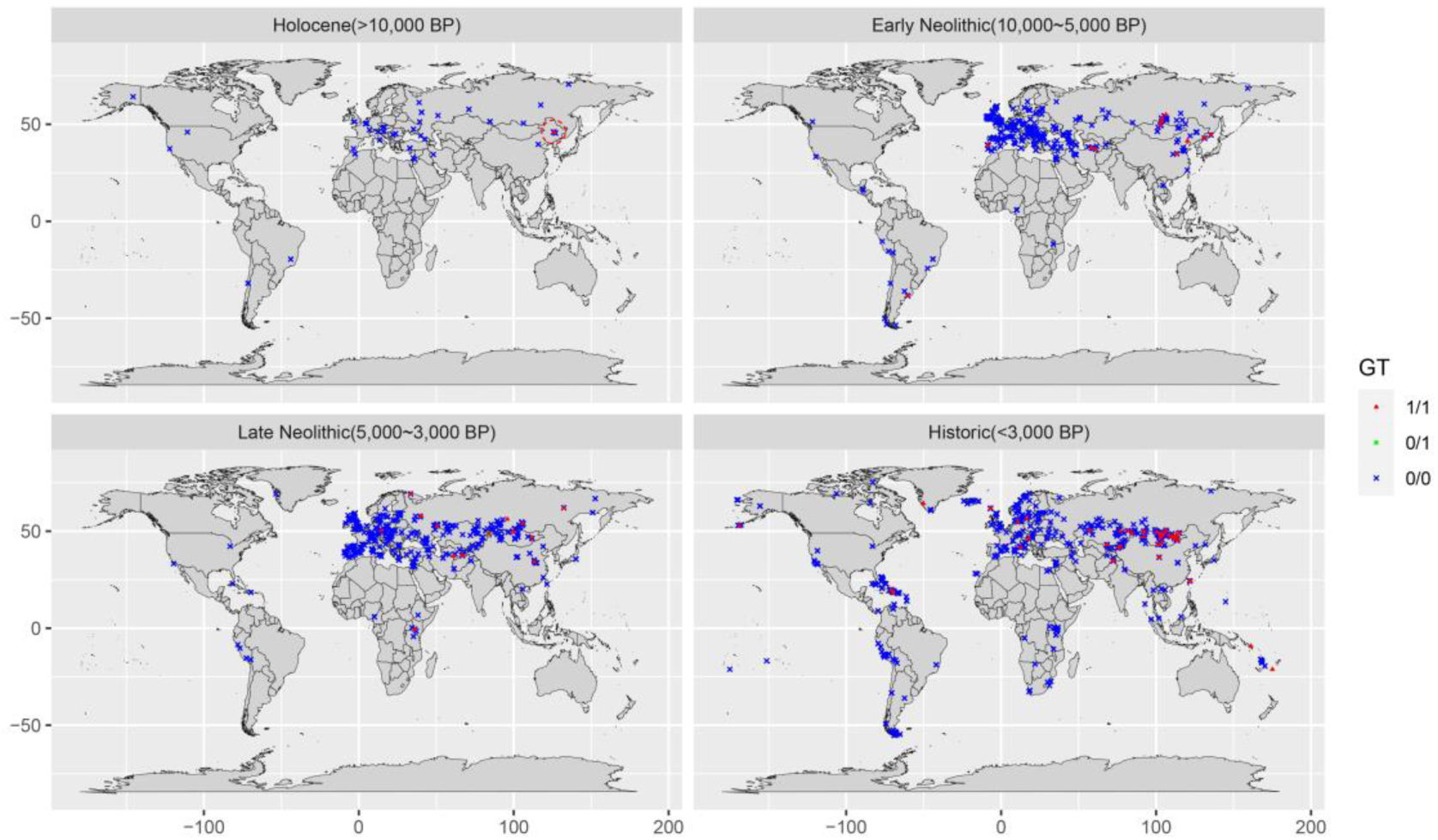
The ancient DNA analysis for SNP rs3781637 *Notes*: the genotype distribution of rs3781637 in ancient populations during four ancient periods.

In addition to *CDKAL1* and *MTNR1B*, the GWAS analysis of the six secondary GDM traits identified other twelve loci, including *HKDC1*, *IGF2BP2*, *PCSK1*, *KCNJ11* (nearest gene), *GLIS3* (nearest gene), *MIR129-1* (nearest gene), *IDE*, *FOXA2*, *NOSTRIN*, *KANK1*, *POLM*, and *ESR1* (Supplementary Figure S4, Table 3). Most of these candidate loci were previously discovered to be associated with pregnancy glycemic traits^31,32^, type II diabetes (T2D) ^12,32^, and serum glucose measurements^33–35^. As a few examples, *HKDC1* encoded a protein that is involved in glucose metabolism and its reduced expression may be associated with GDM, a GWAS study primarily in the European population identified SNP rs4746822 in *HKDC1* associated with 2-hour glucose during OGTT test^32^; *IGF2BP2* plays an important role in metabolism and is susceptible to being associated with diabetes, the Korean study discovered rs1470579 in *IGF2BP2* as a GDM-associated SNP with p-value 2e-07^12^; *KCNJ11* encoded an integral membrane protein and its mutations contribute to neonatal diabetes mellitus and T2D, numerous GWAS studies identified its associations with T2D in both European and East Asian populations^31,36–38^; *IDE* (Insulin Degrading Enzyme) was identified in a European study in 2019^39^, it encodes a zinc metallopeptidase that degrades intracellular insulin, and thereby terminates insulin activity. For the first time, we identified *ESR1* as a candidate glucose-associated gene from GWAS analysis. The gene *ESR1* encodes an estrogen receptor and ligand-activated transcription factor, which regulates many estrogen-inducible genes that play a role in growth, metabolism, sexual development, and gestation^40^. This finding indicates a close relationship between glucose levels and estrogen during pregnancy.

**Table 3.** The associated loci identified from GWAS analysis.

| No. | Loci | CHR | Traits | Summary | EUR |  | EAS |  | CHN |  |
| --- | --- | --- | --- | --- | --- | --- | --- | --- | --- | --- |
|  |  |  |  |  | GDM | Glycemic | GDM | Glycemic | GDM | Glycemic |
| 1 | *CDKALI* | 6 | All pregnancy glycemic | The protein encoded by this gene is a member of the methylthiotransferase family. | X | √ | √ | √ | X | √ |
| 2 | *MTNR1B* | 11 | All pregnancy glycemic | This gene encodes one of two high affinity forms of a receptor for melatonin, the primary hormone secreted by the pineal gland. | X | √ | √ | √ | X | X |
| 3 | *GLIS3* | 9 | OGTT<sub>0h</sub>,<br>OGTT<sub>1h</sub>,<br>OGTT<sub>2h</sub>,<br>GDM<sub>AUC</sub> | This gene is a member of the GLI-similar zinc finger protein family and encodes a nuclear protein with five C2H2-type zinc finger domains. | X | √ | X | √ | X | √ |
| 4 | *PCSK1* | 5 | OGTT<sub>0h</sub> | This gene encodes a member of the subtilisin-like proprotein convertase family | X | √ | X | √ | X | X |
| 5 | *MIR129-1* | 7 | OGTT<sub>1h</sub>,<br>GDM<sub>AUC</sub> | MIR129-1 (MicroRNA 129-1) is an RNA Gene, and is affiliated with the miRNA class. | X | X | X | √ | X | X |
| 6 | *IDE* | 10 | OGTT<sub>1h</sub>,<br>OGTT<sub>1h-0h</sub>,<br>GDM<sub>AUC</sub> | this gene encodes a zinc metalloproteinase that degrades intracellular insulin, and thereby terminates insulins activity | X | √ | X | X | X | X |
| 7 | *FOXA2* | 20 | OGTT<sub>0h</sub> | This gene encodes a member of the forkhead class of DNA-binding proteins. | X | √ | X | √ | X | √ |
| 8 | *NOSTRIN* | 2 | OGTT<sub>0h</sub> | NOSTRIN binds the enzyme responsible for NO production, endothelial NO synthase | X | √ | X | X | X | X |
| 9 | *KANK1* | 9 | OGTT<sub>0h</sub> | The protein encoded by this gene belongs to the Kank family of proteins, which contain multiple ankyrin repeat domains. | X | X | X | √ | X | X |
| 10 | *POLM* | 7 | OGTT<sub>0h</sub> | Predicted to enable DNA-directed DNA polymerase activity. Predicted to be involved in double-strand break repair via nonhomologous end joining. | X | X | X | X | X | X |
| 11 | *ESR1* | 6 | OGTT<sub>0h</sub> | This gene encodes an estrogen receptor and ligand-activated transcription factor. | X | X | X | X | X | X |
| 12 | *IGF2BP2* | 3 | OGTT<sub>1h-0h</sub> | This gene encodes a protein that binds the 5' UTR of insulin-like growth factor 2 (IGF2) mRNA and regulates its translation. | X | √ | √ | √ | X | X |
| 13 | *HKDC1* | 10 | OGTT<sub>2h-0h</sub> | This gene encodes a member of the hexokinase protein family, which is involved in glucose metabolism. Reduced expression may be associated with GDM. | X | √ | X | X | X | X |
| 14 | *KCNJ11* | 11 | OGTT<sub>2h-0h</sub> | The protein encoded by this gene is an integral membrane protein and inward-rectifier type potassium channel. | X | √ | X | √ | X | X |

Notes: CHN is Chinese population.

### Gene-based and functional enrichment analysis

The gene-based results of the principal and secondary GDM traits were provided in Supplementary Table S3. At the threshold of 1e-5, the numbers of significant genes identified for GDM_status_, GDM_risk_, OGTT_0h_, OGTT_1h_, OGTT_2h_, OGTT_1h-0h_, OGTT_2h-0h_, and GDM_AUC_ were 1, 5, 20, 11, 7, 10, 7, and 13. We took a union of these genes and obtained a list of 36 genes, including 27 protein-coding genes, five long-noncoding RNAs (*LINC00261*, *LOC101928266*, *C3orf65*, *LOC101929685*, *CERS6-AS1*), three microRNAs (*MIR129-1*, *MIR4649*, *MIR6884*), and one pseudogene (*PPBPP2*). Twelve out of the fourteen afore-identified loci from single SNP analysis were included in these 36 genes, while two (*GLIS3* and *POLM*) were not. However, the enhancer genes of *GLIS3* and *POLM* were in the 36 genes (*FOXA2* and *GCK*, respectively), showing the GWAS and gene-based analysis results were quite consistent. For the 27 protein-coding genes along with *GLIS3* and *POLM*, we entered them into STRING and obtained their protein-protein interaction (PPI) network and enrichment analysis results (Figure 6a-b, Supplementary Table S4).

**Figure 6.**
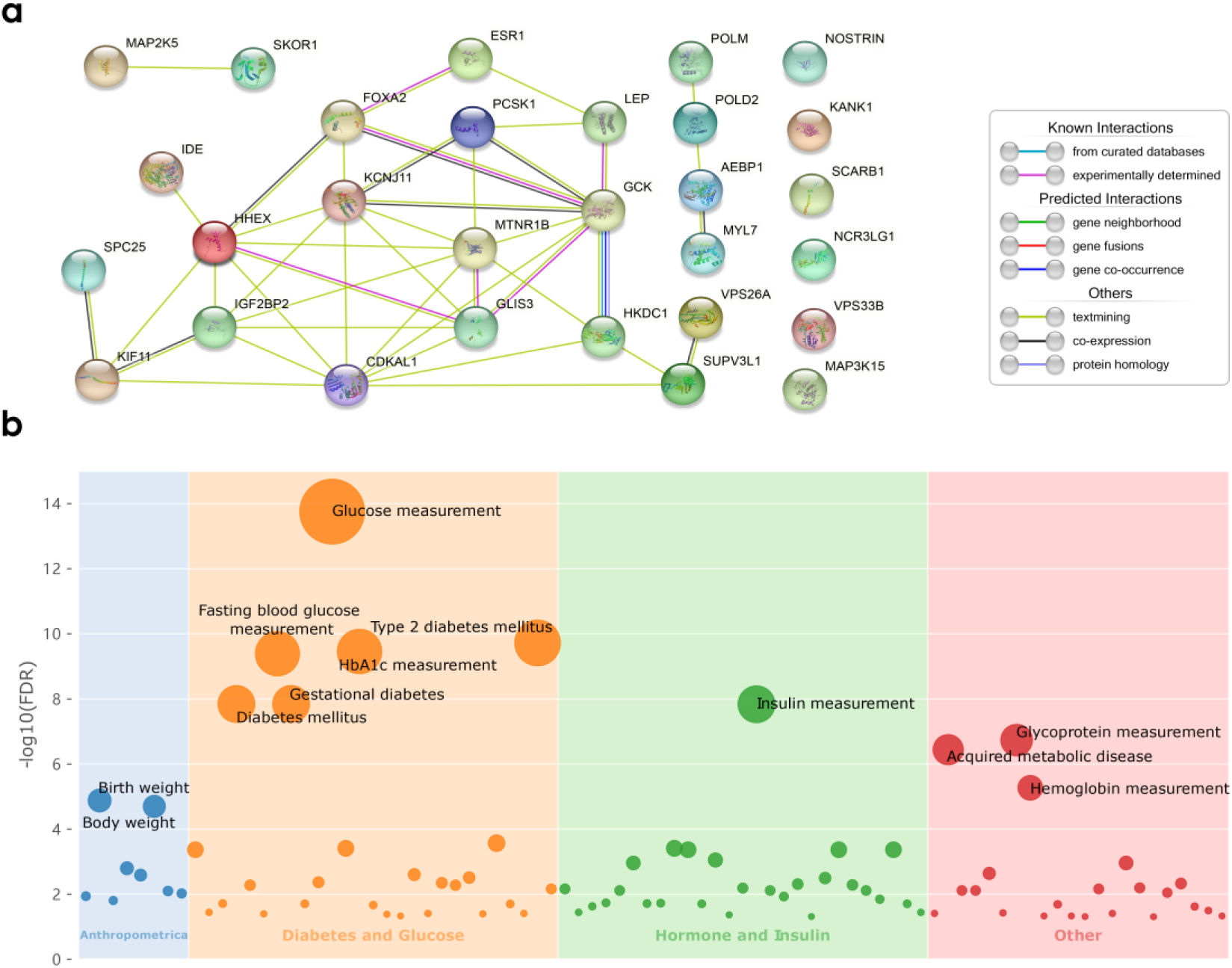
PPI network and results of enrichment analysis *Notes*: **a)** the protein-protein interaction network of candidate genes. **b)** the enriched pathways that passed FDR < 0.05 from the STRING analysis.

The PPI enrichment p-value was < 1.0e-16, meaning the input genes had significant interactions. We recategorized the enriched terms based on their biological functions as: diabetes and glucose (n=15), hormone and insulin (n=16), anthropometric (n=6), and other (n=11). The diabetes and glucose group include glucose measurement (EFO:0004468), type 2 diabetes mellitus (DOID:9352), gestational diabetes (DOID:11714), neonatal diabetes mellitus (DOID:11717), maturity onset diabetes of the young (hsa04950), etc. The hormone and insulin group include insulin measurement (EFO:0004467), regulation of hormone levels (GO:0010817), hormone measurement (EFO:0004730), etc. The anthropometric terms include birth weight (EFO:0004344), body weight (EFO:0004338), body height at birth (EFO:0006784), etc. The other group include glycoprotein measurement (EFO:0004555), acquired metabolic disease (DOID:0060158), parental genotype effect measurement (EFO:0005939), etc.

Many studies have reported the close relationship between GDM and various hormones. Most directly, the hallmark of GDM is that the cells become increasingly resistant to insulin^41^, a hormone that facilitates the transport of glucose from the blood into cells. Other hormones that might contribute to developing GDM include growth hormones, cortisol, estrogen, thyroid, progesterone, placental lactogen, etc. Among them, growth hormones and cortisol are insulin-antagonist hormones, estrogen deficiency may impair glucose homeostasis, lactogen helps break down fat from the mother to provide energy for the fetus, and insulinase inactivates insulin^42,43^. The enrichment results matched what we discovered in the GWAS analysis that hormone levels played an important role in the course of GDM. In addition, the adverse outcomes of GDM, such as neonatal diabetes, high birth weight, childhood obesity, were also revealed from the functional enrichments.

### GDM predictive model construction and evaluation

The construction workflow of the GDM prediction model was given in Figure 7a. Based on the previous analysis, we discovered a series of candidate-associated factors of GDM, including clinical information (women’s age, systolic blood pressure, diastolic blood pressure, weight before pregnancy, and whether first-time pregnancy), lab tests (plasma glucose, γ-glutamyl transferase, prealbumin, urine glucose, and uric acid), and genetic components (two associated SNPs). In this section, we constructed four predictive models: 1) clinical information only, 2) clinical information and early plasma glucose, 3) clinical information and lab tests, and 4) full model: clinical information, lab tests, and genetic components.

**Figure 7.**
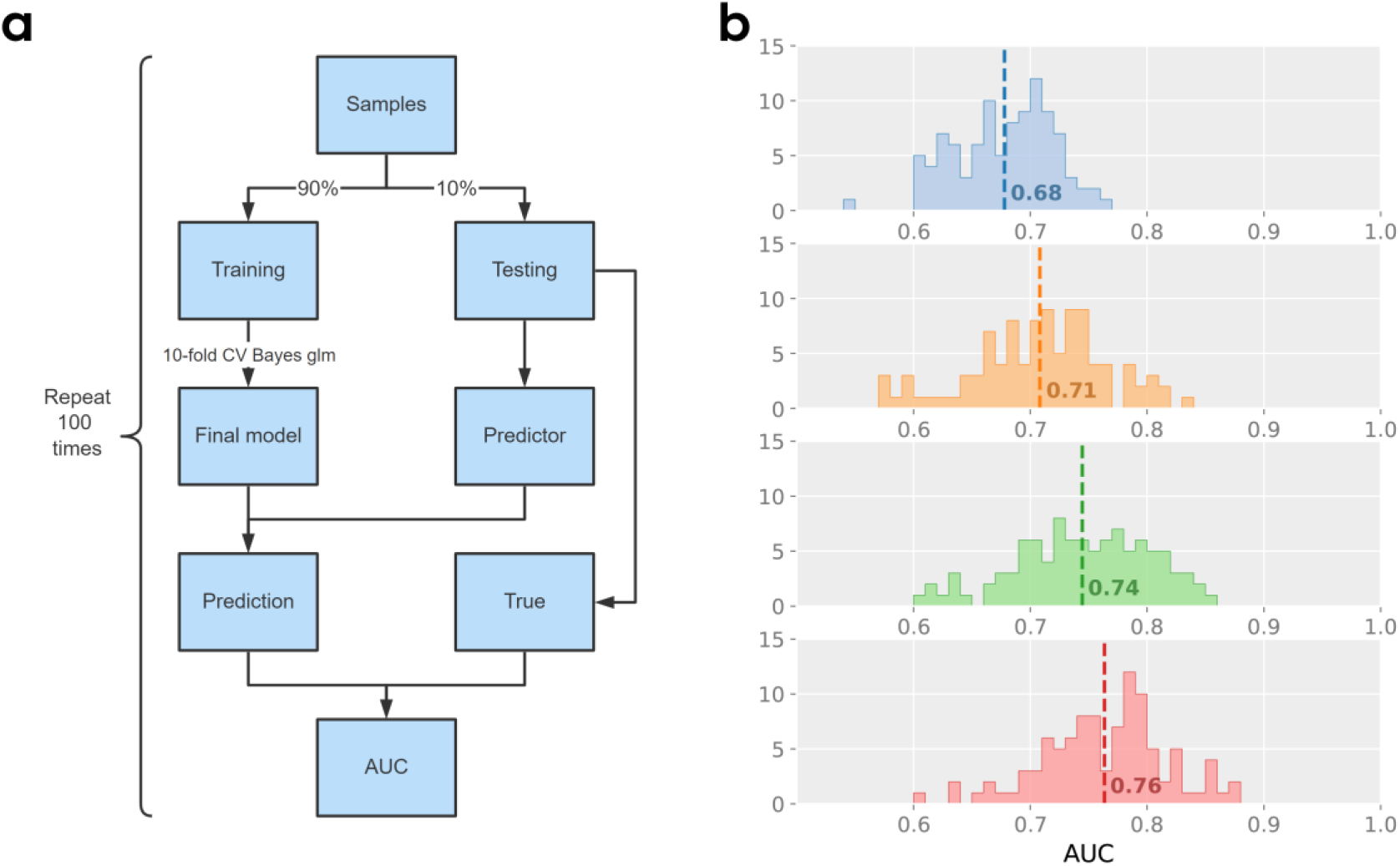
Construction and evaluation of the GDM predictive model *Notes*: **a)** the workflow describes how to construct the predictive model. **b)** the histogram of AUC values under each model. From top to bottom, the models are (1) clinical information only: women’s age, systolic blood pressure, diastolic blood pressure, weight before pregnancy, and whether it’s first-time pregnancy, (2) clinical information and early plasma glucose, (3) clinical information and lab tests: early plasma glucose, γ-glutamyl transferase, prealbumin, urine glucose, and uric acid, (4) full model: clinical information, lab tests, and two SNPs.

We applied histograms to present the distributions of AUC values for each model (Figure 7b). The averaged AUCs for the four models were 0.6779, 0.7082, 0.7445, and 0.7636. After adding glucose levels and other lab tests, the average AUC had a substantial increase, which suggested the necessity of measuring and monitoring plasma glucose and other related factors. Note that, the number of samples who took the glucose test in the early stage was only around 5,000 and thus the sample size in the training set is about 4,500. Some existing predictive models for GDM use only clinical indicators as predictors, with an accuracy of about 65%^44,45^. Some predictive models considered both clinical predictors and genetic factors, but the accuracy is still not greatly improved and is mostly around 70% ^45,46^. Our predictive model uses only ten clinical phenotypes and two SNPs, and the average accuracy is over 76% (median = 0.7715, Q3 = 0.7972) and can exceed 85% in some cases. We also constructed a similar predictive model for GDM_risk_ (Supplementary Figure S5) and the average AUC is 68% for the full model.

## Methods

### Pregnancy glycemic traits

In this study, the subjects were pregnant women who took routine tests during the pregnancy period at Wuhan Children’s Hospital (Wuhan, Hubei Province, Central China) from 2015 to 2020. We excluded seven samples who developed pre-gestational diabetes mellitus (PGDM) prior to pregnancy. The GDM_status_ was diagnosed by oral glucose tolerance test (OGTT) during the 24th to 28th week of pregnancy. The diagnostic criteria were that at least one plasma glucose level in OGTT was reached or exceeded: 5.1 mmol/L (fasting), 10.0 mmol/L (1-hour), and 8.5 mmol/L (2-hour).

To consider the contributions of the three OGTT values simultaneously, we further computed the area under the curve (AUC) as *1-h glucose + (0-h glucose + 2-h glucose)/2* ^20,47^. With the GDM_AUC_ measurements, we further dichotomized all samples into low and high risk of developing GDM (denoted as GDM_risk_). In detail, we defined the high-low risk threshold over a grid of values for GDM_AUC_ while using GDM_status_ as actual values. A ROC curve was then generated for showing the performance at all classification thresholds. Finally, we chose the optimal threshold by optimizing Youden’s J statistics^48^, which determined the best classification threshold and was calculated as the difference between the true positive rate and the false positive rate. In addition, we calculated two differences in OGTT glucose levels, OGTT_1h-0h_ as *1-h glucose* – *0-h glucose* and OGTT_2h-0h_ as *2-h glucose* – *0-h glucose*, for measuring the deterioration of glucose tolerance in 1 and 2 hours^49^.

In the subsequent analysis, we classified all eight pregnancy glycemic traits into two groups: primary traits (GDM_status_, GDM_risk_) and secondary traits (OGTT_0h_, OGTT_1h_, OGTT_2h_, GDM_AUC_, OGTT_1h-0h_, OGTT_2h-0h_. For primary traits, we investigated both environmental and genetic risk factors, the conditional & joint analysis of GWAS results, ancient DNA analysis, and construction of predictive model. For secondary traits, we mainly focused on the genetic contributions by performing GWAS, gene-based analysis and functional protein association analysis.

### Identification of environmental risk factors for primary traits

To determine which clinical information is a potential risk factor of GDM and what adverse postnatal outcomes GDM might cause, we conducted statistical inference on GDM_status_ (GDM_risk_) and available clinical information. For the possible risk factors, we considered pregnant women’s age, weight before pregnancy, BMI, SBP, DBP, and whether first-time pregnancy or multiple pregnancies. The information was collected in the questionnaire survey for the NIPT test. We used BMI > 25 kg/m^2^ as the threshold of obesity. The postnatal outcomes include gestational week, birth weight, newborn gender, and delivery options. Given the gestational week, we defined preterm birth if less than 37 weeks. Low birthweight is when a baby is born weighing less than 2500 g, and a baby who weighs more than 4000 g at birth is diagnosed as having fetal macrosomia (a.k.a. giant baby). The delivery options include natural birth and cesarean section (c-section). If the clinical variable is quantitative, we used the two-sample t-test to determine if the variable means are equal between case and control groups. Otherwise, we used the chi-squared test to check whether GDM_status_ or GDM_risk_ is likely to be related to the binary variable.

In addition, a wide spectrum of laboratory tests was collected, such as serum and urine biomarkers (e.g., serum glucose, liver function, urine acid). We used the first 15 gestational weeks as the first trimester and assembled the maternal tests performed in this period. For continuous traits, the mean value was taken for multiple examinations; for multiple dichotomous traits, a positive status was defined once a positive result occurred. To identify possibly associated laboratory measurements of GDM, we performed logistic regression analysis. For the logistic regression model, we used women’s age as a covariate, the GDM_status_ and GDM_risk_ as response variables, and the laboratory traits as predictors one at a time.

### Genome-wide association study

In this section, we performed GWAS analysis for the pregnancy glycemic traits. The genotype data processing procedure was the same as in the companion work [cite]. The GWAS analysis in PLINK 2.0^50^ with argument *--glm - -maf 0.05 --hwe 1e-06 --geno 0.01*. The logistic and linear regression models were applied for primary and secondary traits, respectively. The covariates included pregnant women’s age and five principal components (PCs) of genotype variants. All covariates were standardized to have zero mean and one standard deviation by using the argument *--covar-variance-standardize* and all quantitative phenotypes were normalized by *-- pheno-quantile-normalize*. We set the genome-wide significance threshold as 5e-08. Based on the GWAS results of the primary traits, we further performed conditional and joint association analysis in GCTA^51,52^ for identifying independent significant signals. The LD correlations were calculated from the 1000 genome phase 3 database.

### Gene-based and functional enrichment analysis

Based on the GWAS summary statistics of each glycemic trait, we performed gene- and pathway-based association analysis to identify candidate genes and biological pathways. First, to do gene-based testing, we used the online server of VEGAS2 (https://vegas2.qimrberghofer.edu.au/)^53^ by selecting Asian as the reference population and a window of size 50 kb outside each gene. Further, we took a union of significant genes associated with each trait at a threshold of 1e-5. We then used STRING (https://string-db.org) to get the protein-protein interaction (PPI) network to visualize the interactions of these candidate genes^54^. Finally, based on the list of genes/proteins, STRING could highlight functional enrichments using a number of functional classification systems such as Gene Ontology (GO), Kyoto Encyclopedia of Genes and Genomes (KEGG), Disease-gene associations (Diseases), and Human Phenotype Ontology (Monarch HPO).

### GDM predictive model construction and evaluation

Based on the environmental and genetic risk factors, we constructed predictive models for GDM_status_ and GDM_risk_. We considered several models: 1) risk clinical factors only, 2) clinical factors and laboratory measurements, and 3) clinical factors, laboratory measurements, and genetic factors. For each model, we randomly classified all samples into a training set (90%) and a testing test (10%). For the training set, we performed ten-fold cross-validation to train the model and used Bayes generalization linear model to estimate the model coefficients. This was achieved in R software with function *caret:train*. After obtaining the final model, we multiplied the produced coefficients by predictors in the testing set, which were the prediction values. Finally, we calculated the AUC (area under the curve) for prediction and true values in R with function *pROC:auc*. We replicated the entire procedure 100 times and computed a mean AUC. The model construction workflow was given in Figure 7a.

## Discussion

Gestational diabetes mellitus (GDM) is one of the common complications of pregnancy, and there are few studies on its genetic background in the Chinese population, and most of the available studies are limited by sample size. In this study, using maternal genotype data obtained from high-throughput noninvasive prenatal genetic testing, we conducted the hitherto largest GWAS analysis of GDM in Asian population and identified rs35261542, rs4712530, rs3781637, and rs12225378. The allele frequencies of the latter three variants are substantially different in European and East Asian populations, which might suggest distinct genetic background of GDM in the two populations. To be highlighted, the SNP rs3781637 was originally mutated in an ancient Chinese over 10,000 years ago and had higher frequency in EAS than EUR at present. The gene set enrichment analysis revealed functional pathways, including diabetes and glucose, hormone and insulin, and intriguingly neonatal phenotypes, such as neonatal diabetes mellitus and birth weight. Our predictive model with both environmental and genetic factors as predictors reached an acceptable accuracy for early GDM prediction.

Although our work provided several insights into the genetic etiology of GDM, it also had several shortcomings. First, even though the sample size we used was by far the largest in the Asian population and we uncovered numerous novel loci associated with pregnancy glycemic traits, specifically for GDM_status_ (GDM_risk_), we only replicated two associated loci, *MTNR1B* and *CDKAL1*. We are currently recruiting more samples and aim to perform a larger-scale GWAS or meta-analysis to discover more candidate genes. Second, although the mean AUC of our predictive model was over 0.76, it is still a moderately acceptable performance. There are two reasons for this: the number of samples who took glucose tests in the first trimester was very small (around 4,000), which resulted in small sizes of training and testing sets; and the genotype data were ultra-low depth sequencing data, and although genotype imputation accuracy was high and could meet the requirements of subsequent population-level genetic studies, there was still less certainty for individual genotype data. The future efforts may focus on two aspects: first, to further improve the genotype imputation accuracy to obtain more accurate individual genotype data; and second, to use large-scale low-depth sequencing data as an exploration dataset for mining GDM-associated loci, and then use high-depth sequencing data as a prediction dataset to construct predictive models.

Gestational diabetes mellitus is a very complex disease of pregnancy and its pathogenesis is still not well understood. Our study explored the loci and phenotypes associated with GDM at the genetic and clinical levels and constructed a predictive model accordingly, which is an important reference for revealing the pathogenetic mechanism of GDM.

## Data Availability

All data produced in the present study are available upon reasonable request to the authors

## Author contribution

A.Z. and X.J. conceived the study, designed the research program, and managed the project.

H.X., M.Y., J.Z., Y.Z., J.L., Y.Y.Z., Z.C., H.M., X.C., L.H., and R.Z. collected the data.

L.L., X.L., and P.L. preprocessed the data and finished the quality control.

H.Z., H.X., L.L., M.Y., M.C., X.Z, X.Q, and J.Y.Z. performed the statistical analysis and results visualization.

H.Z., H.X., L.L., M.Y., and M.C. wrote the manuscript.

All authors participated in revising the manuscript.

## Acknowledgment

This study was supported by the Central Guidance on Local Science and Technology Development Fund of Hubei Province (2022BGE261), National Natural Science Foundation of China (32171441 and 32000398), Guangdong-Hong Kong Joint Laboratory on Immunological and Genetic Kidney Diseases (2019B121205005), Top Medical Young Talents (2019) of Hubei Province, Guangdong Provincial Key Laboratory of Genome Read and Write (2017B030301011), open project of BGI-Shenzhen, Shenzhen 518000 China (BGIRSZ20200008) and the China National GeneBank.

## Data availability

The GWAS summary statistics of all eight pregnancy glycemic traits have been deposited into CNGB Sequence Archive (CNSA)^55^ of China National GeneBank DataBase (CNGBdb)^56^ with accession number CNP0003674.

## Supplementary Figures

**Supplementary Figure S1.**
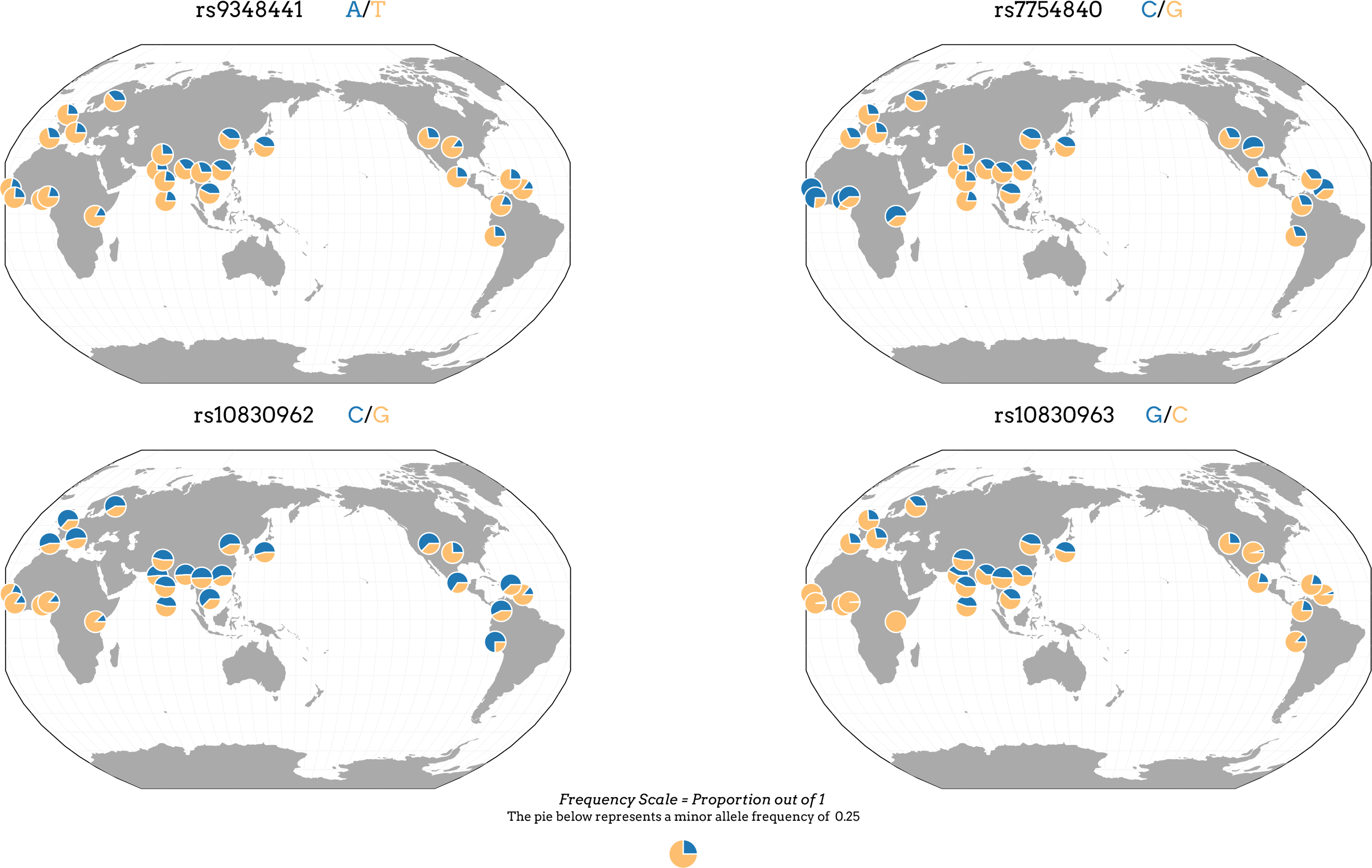
The global geographic distributions of additional SNPs

**Supplementary Figure S2.**
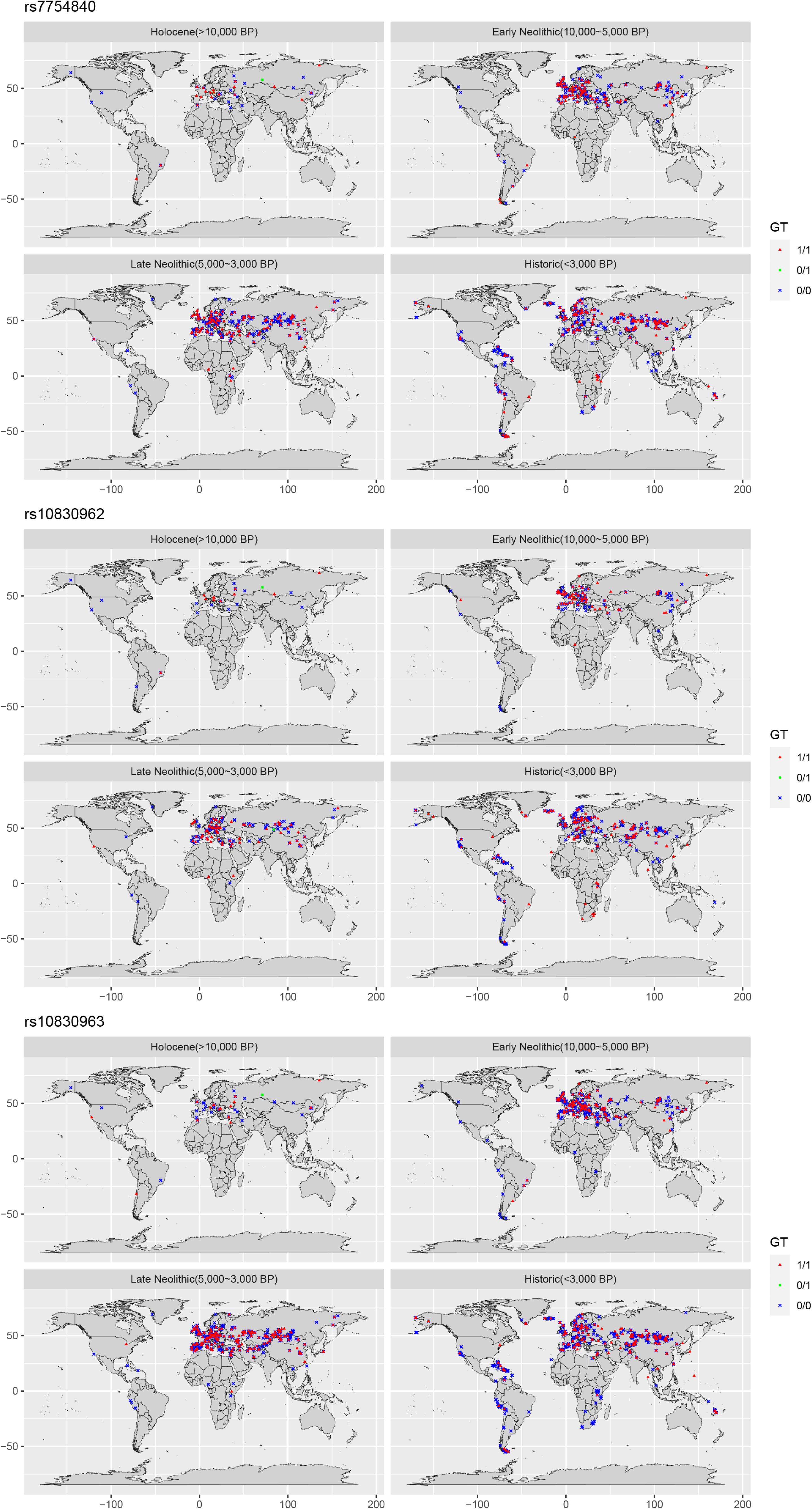
The ancient DNA analysis for additional SNPs

**Supplementary Figure S3.**
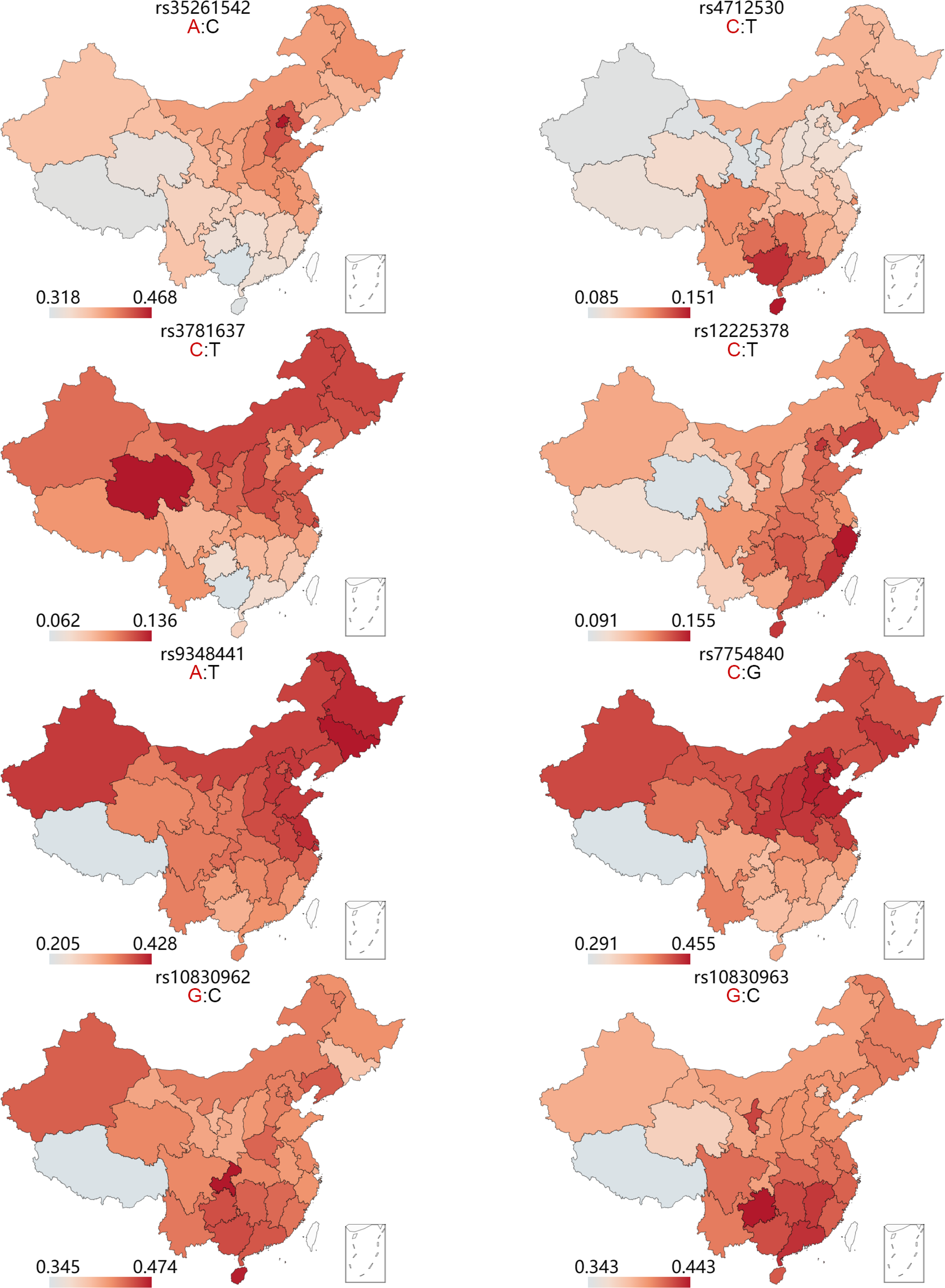
The allele frequency distributions in different provinces of China for identified and additional SNPs

**Supplementary Figure S4.**
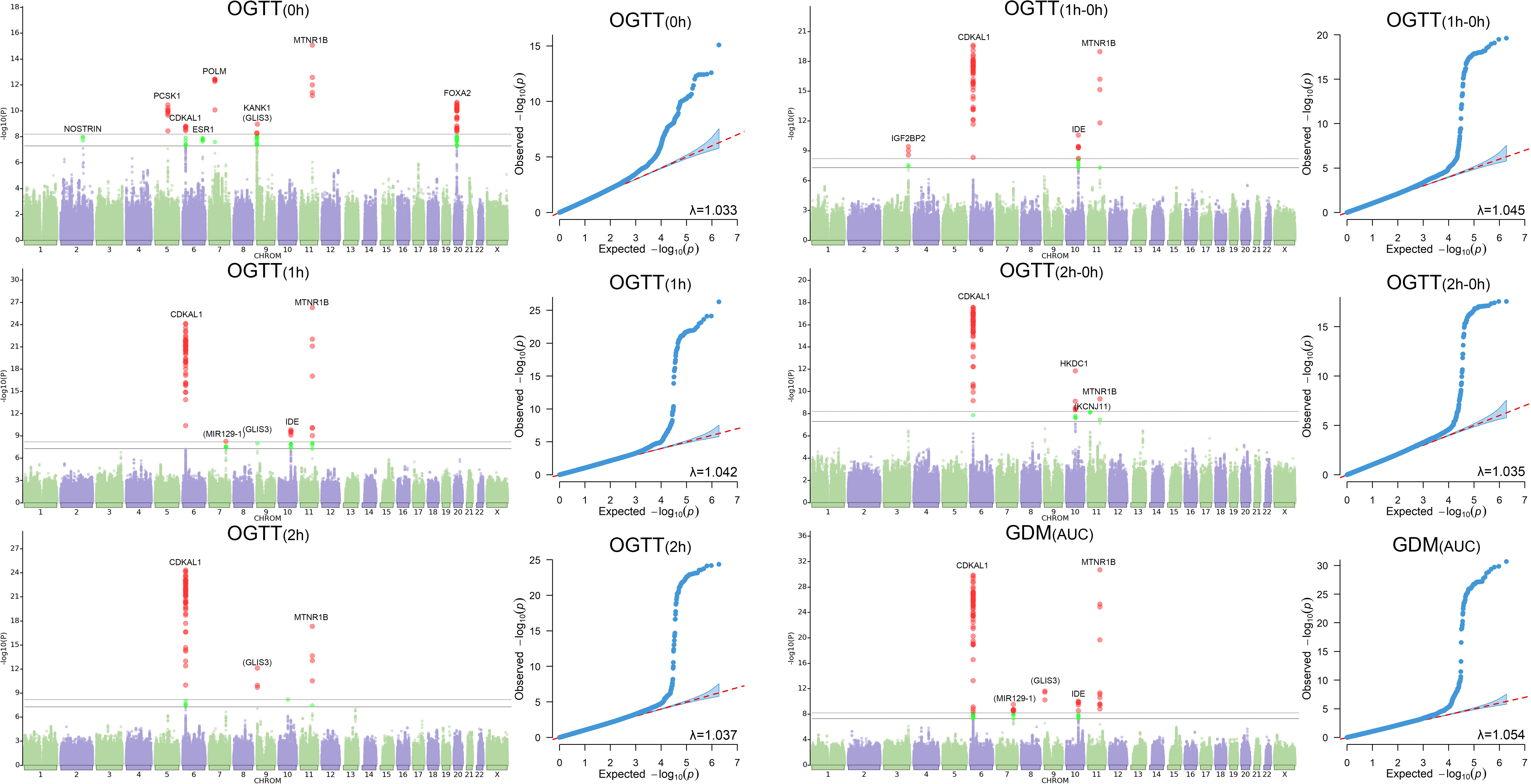
The GWAS results of the secondary GDM traits

**Supplementary Figure S5.**
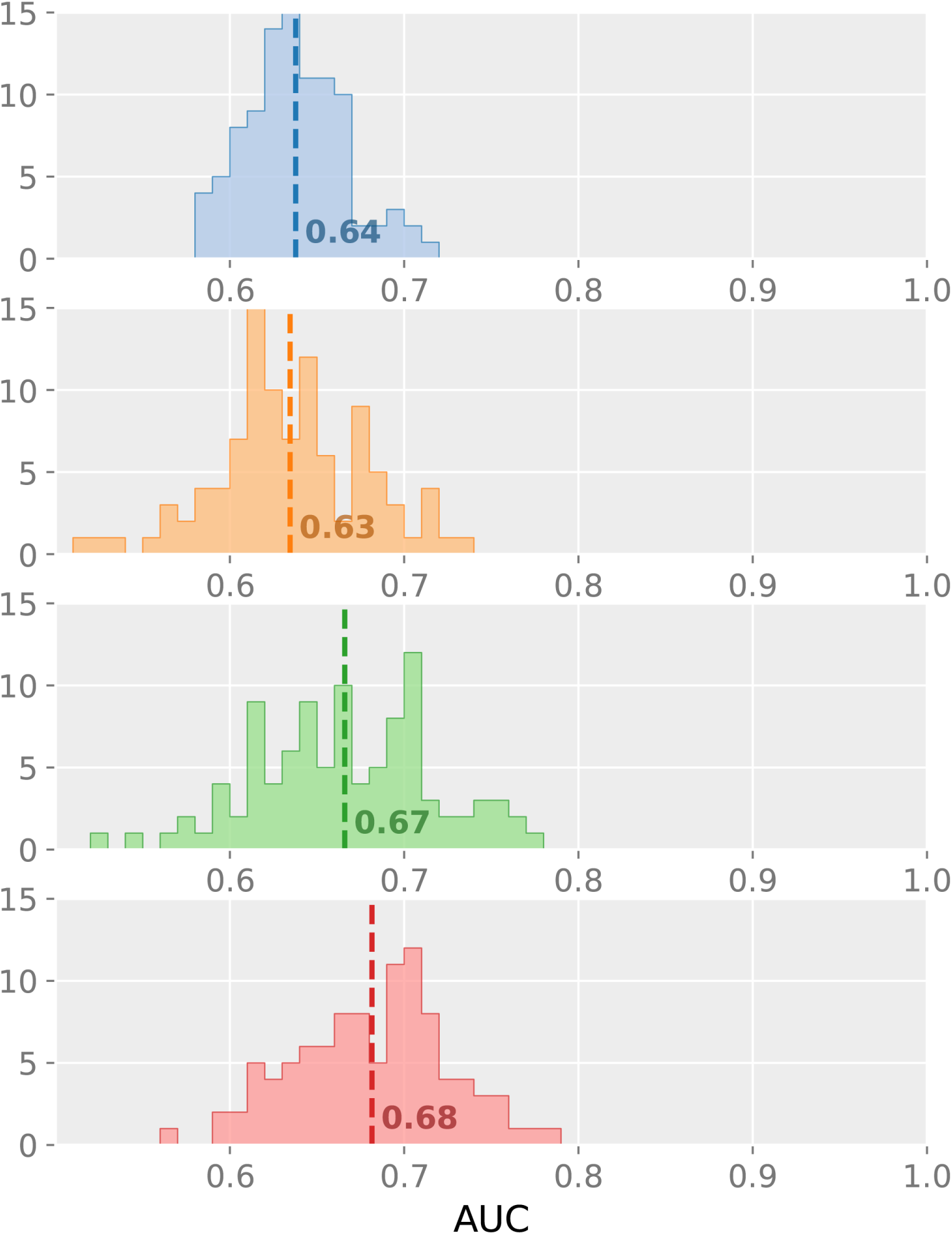
The histogram of AUC values for prediction model of GDM_risk_

**Table S1.**
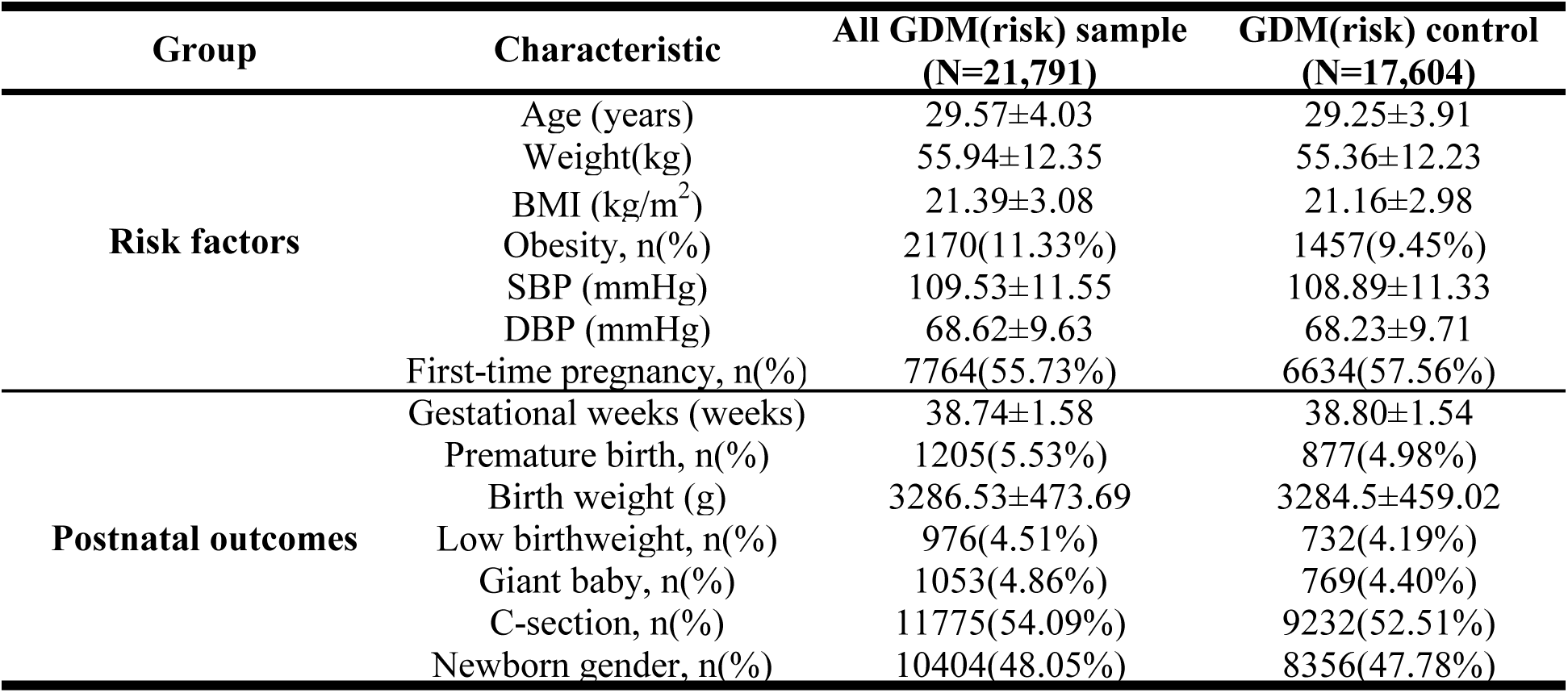
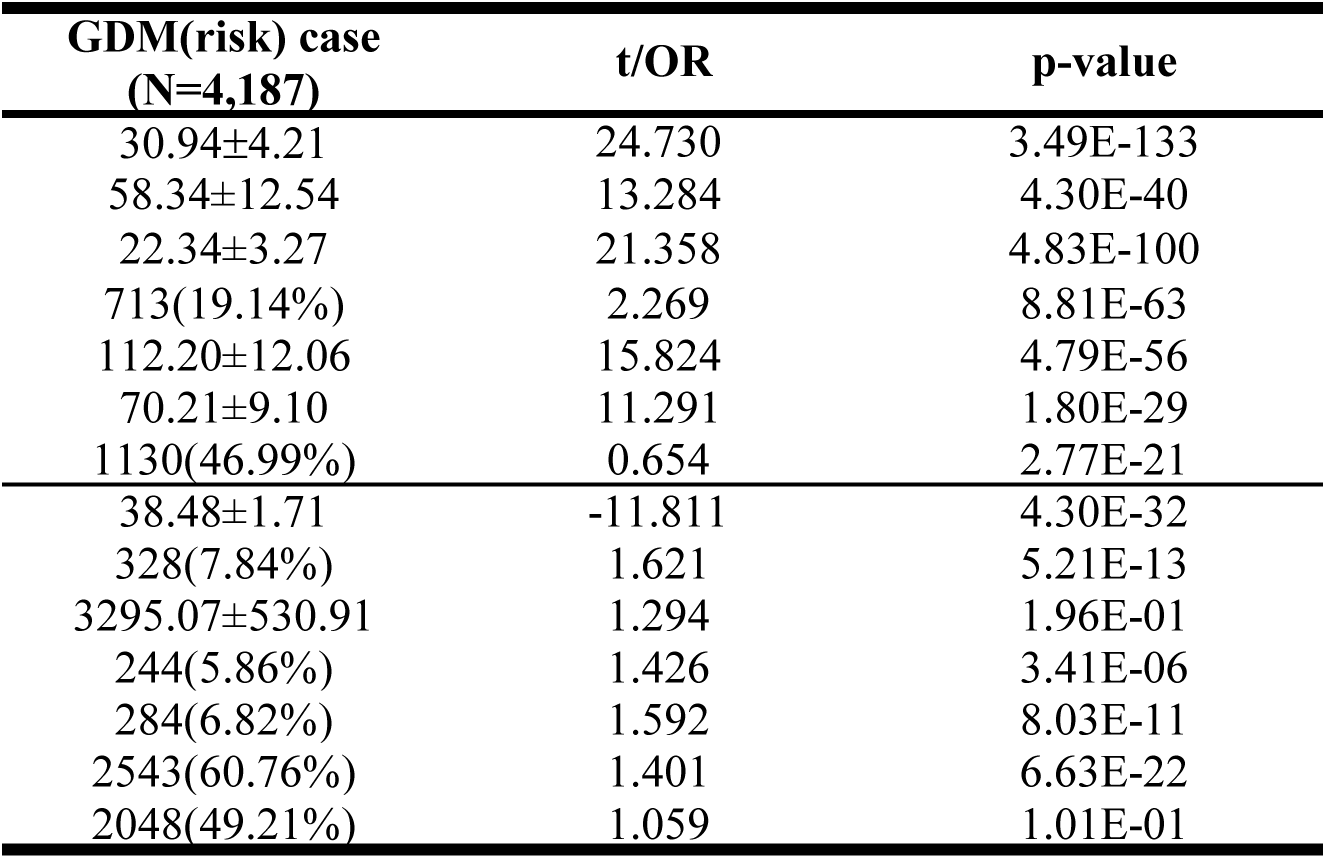
Regression analysis between clinical information and GDM(risk)

**Table S2.**
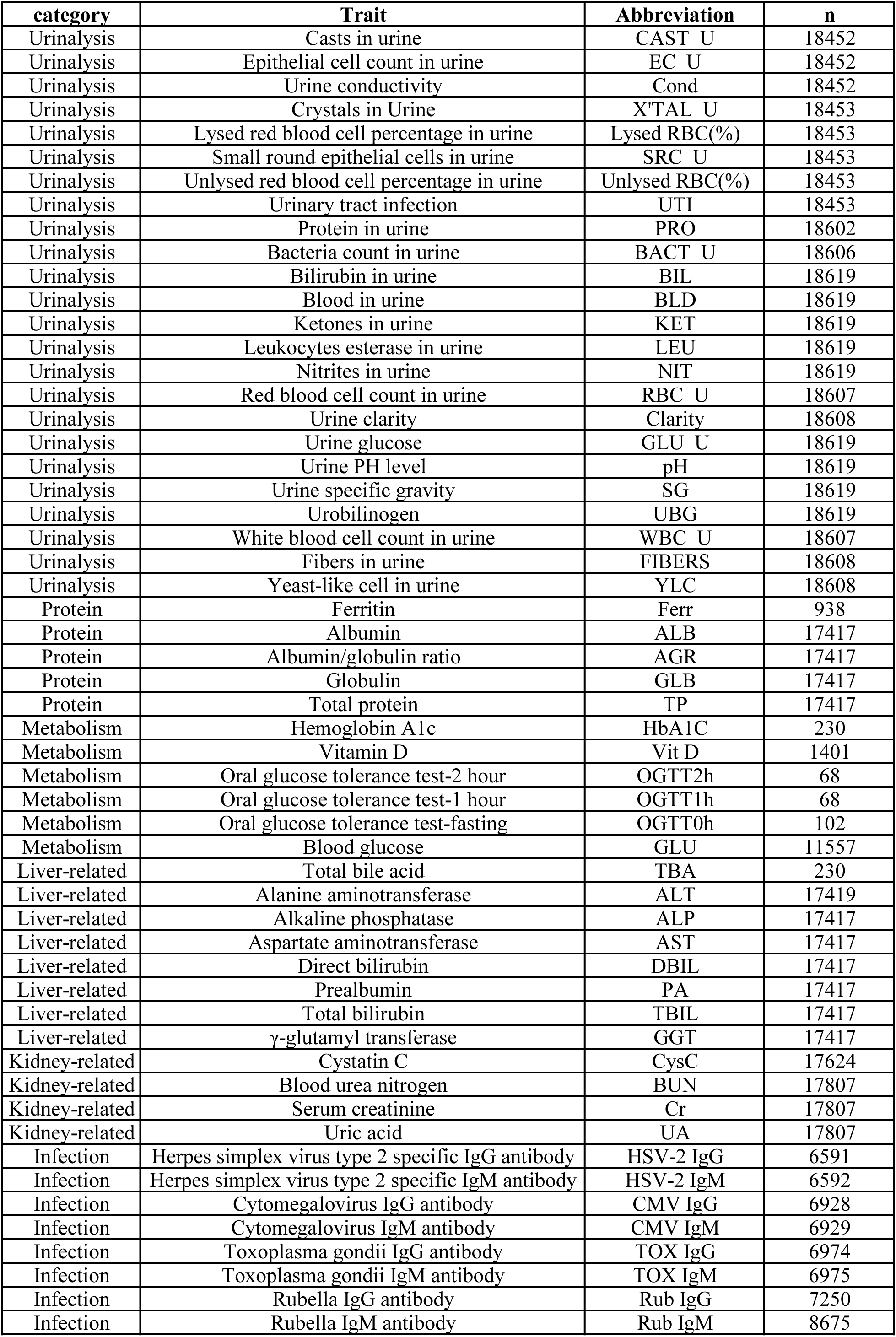
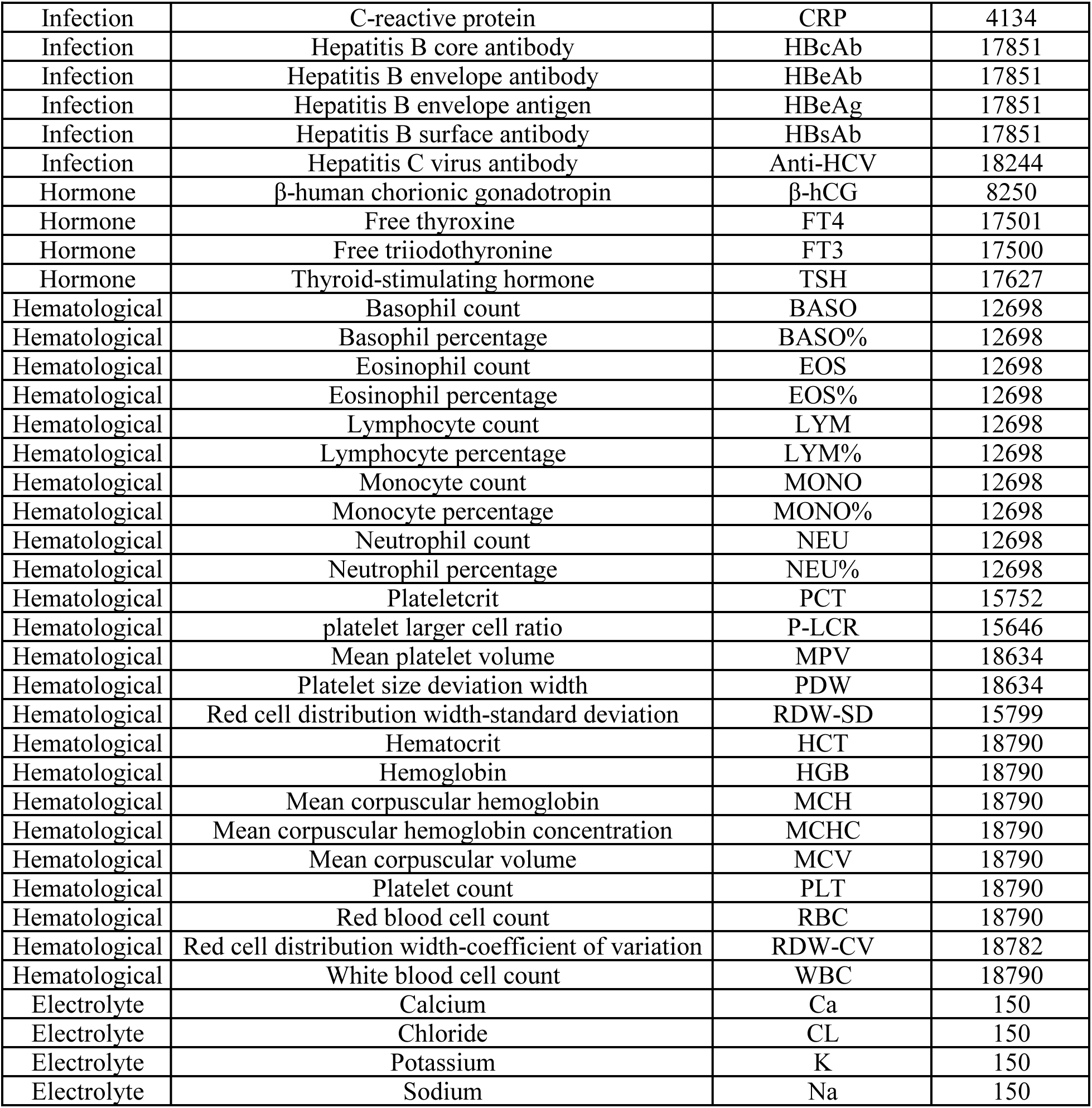
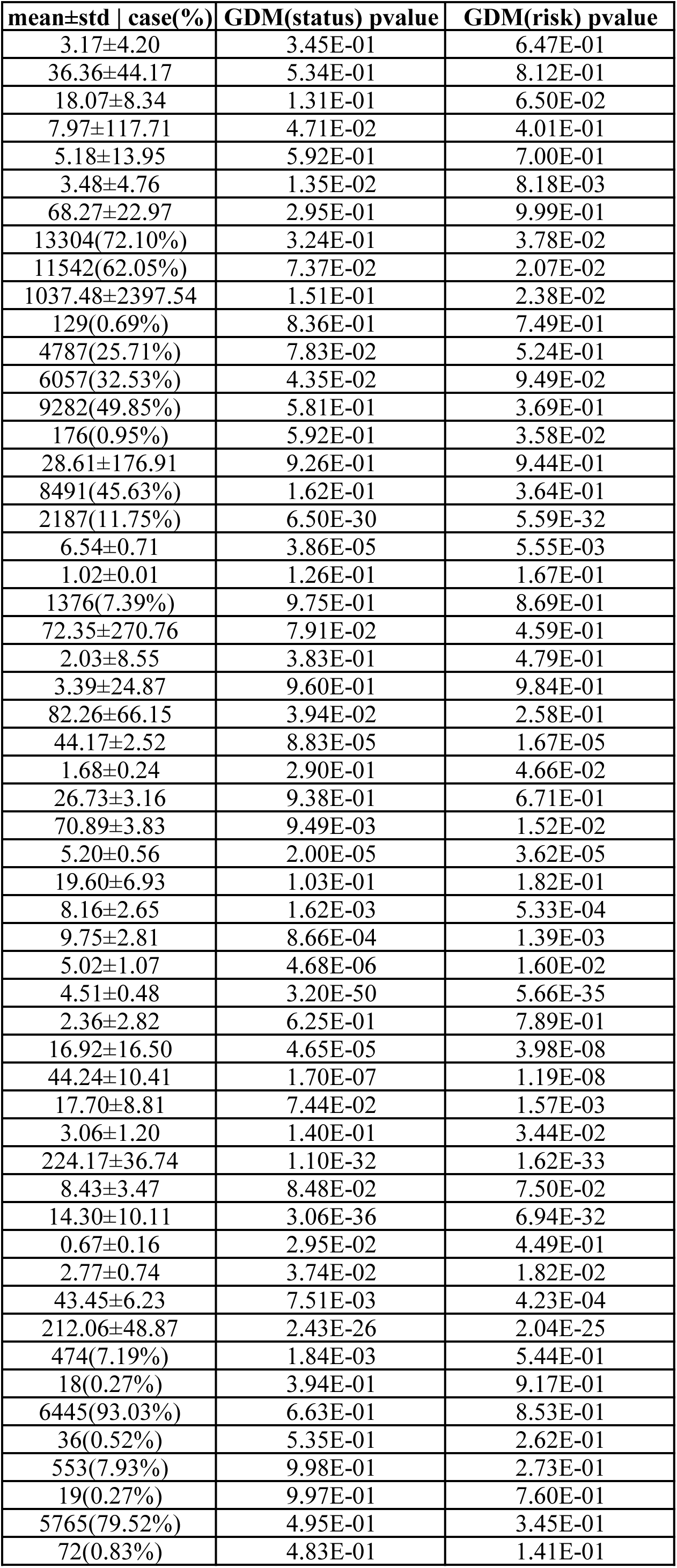
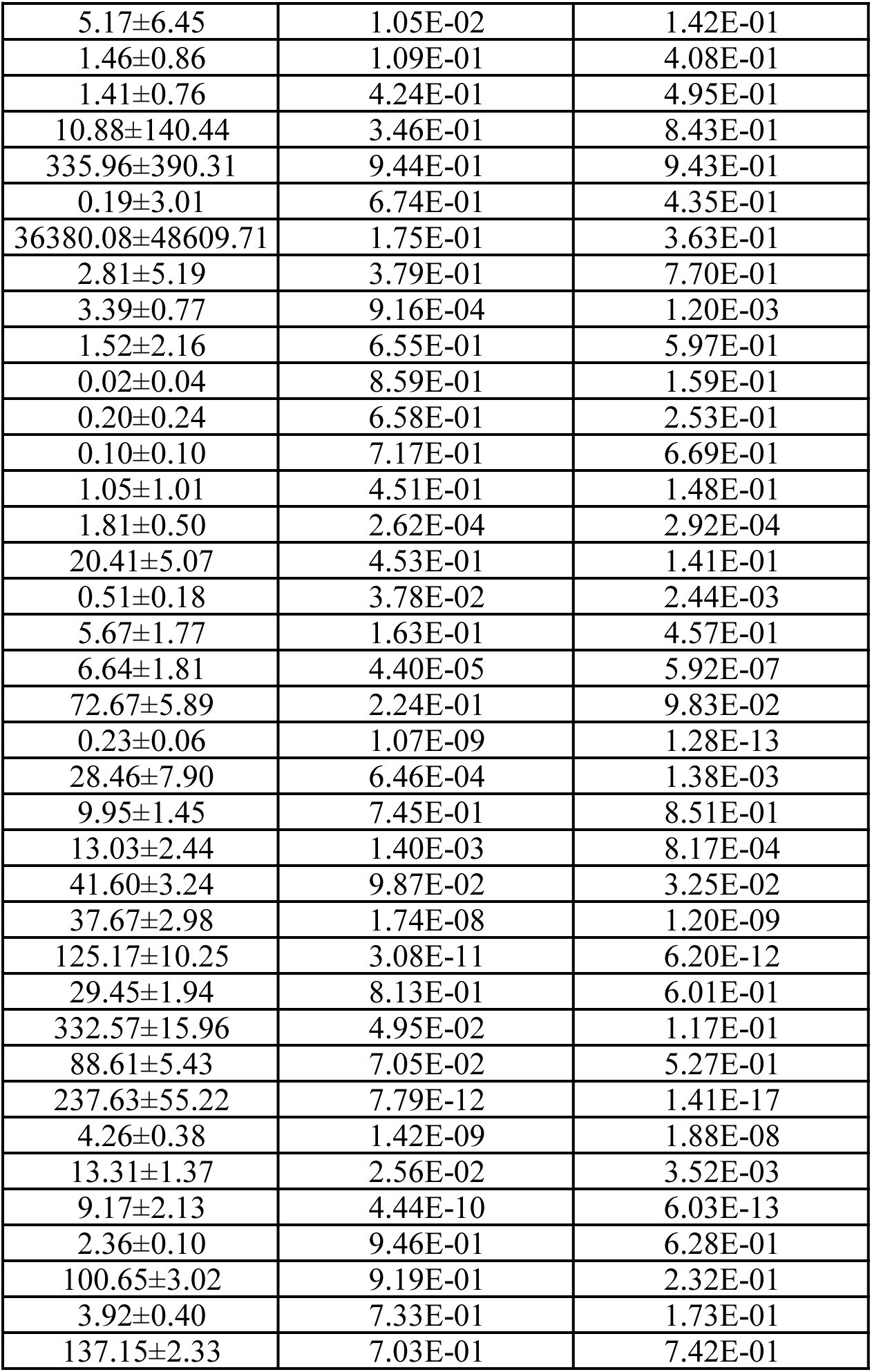
The regression analysis results of the principal GDM traits on laboratory traits in the first trimester.

**Table S3.** The list of identified genes from gene-based association analysis for all pregnancy glycemic traits.

| trait | Gene | nSNPs | nSims | Chr | Start | Stop | Test |
| --- | --- | --- | --- | --- | --- | --- | --- |
| GDM | CDKAL1 | 534 | 1.00E+06 | 6 | 20484687 | 21282634 | 3675.151 |
| GDM(AUC) | IDE | 143 | 1.00E+06 | 10 | 94161440 | 94383852 | 1410.67 |
| GDM(AUC) | KIF11 | 147 | 1.00E+06 | 10 | 94302824 | 94465152 | 1742.978 |
| GDM(AUC) | MTNR1B | 29 | 1.00E+06 | 11 | 92652788 | 92765948 | 753.406 |
| GDM(AUC) | CDKAL1 | 534 | 1.00E+06 | 6 | 20484687 | 21282634 | 10106.36 |
| GDM(AUC) | LEP | 56 | 1.00E+06 | 7 | 127831330 | 127947682 | 720.9331 |
| GDM(AUC) | MIR129-1 | 46 | 1.00E+06 | 7 | 127797924 | 127897996 | 691.9856 |
| GDM(AUC) | HHEX | 57 | 1.00E+06 | 10 | 94399680 | 94505408 | 682.1587 |
| GDM(AUC) | IGF2BP2 | 77 | 1.00E+06 | 3 | 185311526 | 185592827 | 611.4707 |
| GDM(AUC) | MYL7 | 21 | 1.00E+06 | 7 | 44128462 | 44230916 | 164.459 |
| GDM(AUC) | SKOR1 | 135 | 1.00E+06 | 15 | 68062041 | 68176174 | 1058.758 |
| GDM(AUC) | MAP3K15 | 52 | 1.00E+06 | 23 | 19328175 | 19583379 | 481.4499 |
| GDM(AUC) | MAP2K5 | 345 | 1.00E+06 | 15 | 67785020 | 68149455 | 2787.699 |
| GDM(AUC) | LINC00261 | 69 | 1.00E+06 | 20 | 22491191 | 22609280 | 834.9323 |
| GDM(risk) | MTNR1B | 29 | 1.00E+06 | 11 | 92652788 | 92765948 | 453.6538 |
| GDM(risk) | CDKAL1 | 534 | 1.00E+06 | 6 | 20484687 | 21282634 | 4547.797 |
| GDM(risk) | LEP | 56 | 1.00E+06 | 7 | 127831330 | 127947682 | 479.188 |
| GDM(risk) | MIR129-1 | 46 | 1.00E+06 | 7 | 127797924 | 127897996 | 457.0015 |
| OGTT(1h-0h) | IDE | 143 | 1.00E+06 | 10 | 94161440 | 94383852 | 1537.928 |
| OGTT(1h-0h) | MTNR1B | 29 | 1.00E+06 | 11 | 92652788 | 92765948 | 443.5011 |
| OGTT(1h-0h) | IGF2BP2 | 77 | 1.00E+06 | 3 | 185311526 | 185592827 | 973.2522 |
| OGTT(1h-0h) | CDKAL1 | 534 | 1.00E+06 | 6 | 20484687 | 21282634 | 6631.721 |
| OGTT(1h-0h) | LEP | 56 | 1.00E+06 | 7 | 127831330 | 127947682 | 435.937 |
| OGTT(1h-0h) | MIR129-1 | 46 | 1.00E+06 | 7 | 127797924 | 127897996 | 409.0499 |
| OGTT(1h-0h) | KIF11 | 147 | 1.00E+06 | 10 | 94302824 | 94465152 | 1729.682 |
| OGTT(1h-0h) | LOC101928266 | 44 | 1.00E+06 | 17 | 9024384 | 9132435 | 234.0735 |
| OGTT(1h-0h) | HHEX | 57 | 1.00E+06 | 10 | 94399680 | 94505408 | 651.2143 |
| OGTT(1h-0h) | C3orf65 | 13 | 1.00E+06 | 3 | 185381039 | 185485955 | 99.64819 |
| OGTT(2h-0h) | HKDC1 | 89 | 1.00E+06 | 10 | 70930058 | 71077315 | 678.9125 |
| OGTT(2h-0h) | SUPV3L1 | 66 | 1.00E+06 | 10 | 70889992 | 71018849 | 759.2186 |
| OGTT(2h-0h) | NCR3LG1 | 56 | 1.00E+06 | 11 | 17323308 | 17448868 | 578.3214 |
| OGTT(2h-0h) | IGF2BP2 | 77 | 1.00E+06 | 3 | 185311526 | 185592827 | 696.7233 |
| OGTT(2h-0h) | CDKAL1 | 534 | 1.00E+06 | 6 | 20484687 | 21282634 | 5685.58 |
| OGTT(2h-0h) | VPS26A | 63 | 1.00E+06 | 10 | 70833907 | 70982616 | 665.0363 |
| OGTT(2h-0h) | KCNJ11 | 48 | 1.00E+06 | 11 | 17356795 | 17460878 | 387.2986 |
| OGTT(0h) | MTNR1B | 29 | 1.00E+06 | 11 | 92652788 | 92765948 | 468.9298 |
| OGTT(0h) | FOXA2 | 66 | 1.00E+06 | 20 | 22511641 | 22616101 | 2002.799 |
| OGTT(0h) | LINC00261 | 69 | 1.00E+06 | 20 | 22491191 | 22609280 | 2038.685 |
| OGTT(0h) | LOC101929685 | 79 | 1.00E+06 | 20 | 22518159 | 22638155 | 2372.763 |
| OGTT(0h) | NOSTRIN | 49 | 1.00E+06 | 2 | 169593048 | 169771849 | 448.3054 |
| OGTT(0h) | PCSK1 | 61 | 1.00E+06 | 5 | 95676039 | 95818985 | 883.8252 |
| OGTT(0h) | CDKAL1 | 534 | 1.00E+06 | 6 | 20484687 | 21282634 | 3374.808 |
| OGTT(0h) | AEBP1 | 26 | 1.00E+06 | 7 | 44093959 | 44204164 | 440.5123 |
| OGTT(0h) | GCK | 47 | 1.00E+06 | 7 | 44133869 | 44279022 | 795.9898 |
| OGTT(0h) | LEP | 56 | 1.00E+06 | 7 | 127831330 | 127947682 | 427.7194 |
| OGTT(0h) | MIR129-1 | 46 | 1.00E+06 | 7 | 127797924 | 127897996 | 446.5559 |
| OGTT(0h) | MIR4649 | 24 | 1.00E+06 | 7 | 44100447 | 44200511 | 439.564 |
| OGTT(0h) | MYL7 | 21 | 1.00E+06 | 7 | 44128462 | 44230916 | 483.3359 |
| OGTT(0h) | POLD2 | 23 | 1.00E+06 | 7 | 44104278 | 44213169 | 433.7268 |
| OGTT(0h) | KANK1 | 410 | 1.00E+06 | 9 | 420293 | 796106 | 3755.757 |
| OGTT(0h) | SPC25 | 8 | 1.00E+06 | 2 | 169677400 | 169796944 | 87.23131 |
| OGTT(0h) | CERS6-AS1 | 57 | 1.00E+06 | 2 | 169578460 | 169692939 | 463.285 |
| OGTT(0h) | SCARB1 | 82 | 1.00E+06 | 12 | 125212173 | 125398519 | 365.4025 |
| OGTT(0h) | ESR1 | 368 | 1.00E+06 | 6 | 151961630 | 152474408 | 2698.664 |
| OGTT(0h) | MIR6884 | 63 | 1.00E+06 | 17 | 38132584 | 38232662 | 640.0471 |
| OGTT(1h) | IDE | 143 | 1.00E+06 | 10 | 94161440 | 94383852 | 1501.187 |
| OGTT(1h) | KIF11 | 147 | 1.00E+06 | 10 | 94302824 | 94465152 | 1709.808 |
| OGTT(1h) | MTNR1B | 29 | 1.00E+06 | 11 | 92652788 | 92765948 | 658.0288 |
| OGTT(1h) | LOC101928266 | 44 | 1.00E+06 | 17 | 9024384 | 9132435 | 250.667 |
| OGTT(1h) | IGF2BP2 | 77 | 1.00E+06 | 3 | 185311526 | 185592827 | 682.8723 |
| OGTT(1h) | CDKAL1 | 534 | 1.00E+06 | 6 | 20484687 | 21282634 | 8194.91 |
| OGTT(1h) | LEP | 56 | 1.00E+06 | 7 | 127831330 | 127947682 | 637.0935 |
| OGTT(1h) | MIR129-1 | 46 | 1.00E+06 | 7 | 127797924 | 127897996 | 616.1683 |
| OGTT(1h) | HHEX | 57 | 1.00E+06 | 10 | 94399680 | 94505408 | 648.2479 |
| OGTT(1h) | MAP2K5 | 345 | 1.00E+06 | 15 | 67785020 | 68149455 | 2680.542 |
| OGTT(1h) | PPBPP2 | 9 | 1.00E+06 | 4 | 74869754 | 74971116 | 57.3616 |
| OGTT(2h) | SUPV3L1 | 66 | 1.00E+06 | 10 | 70889992 | 71018849 | 512.9142 |
| OGTT(2h) | MTNR1B | 29 | 1.00E+06 | 11 | 92652788 | 92765948 | 383.9974 |
| OGTT(2h) | CDKAL1 | 534 | 1.00E+06 | 6 | 20484687 | 21282634 | 8088.592 |
| OGTT(2h) | LEP | 56 | 1.00E+06 | 7 | 127831330 | 127947682 | 480.615 |
| OGTT(2h) | MIR129-1 | 46 | 1.00E+06 | 7 | 127797924 | 127897996 | 431.8365 |
| OGTT(2h) | HKDC1 | 89 | 1.00E+06 | 10 | 70930058 | 71077315 | 476.4645 |
| OGTT(2h) | PCSK1 | 61 | 1.00E+06 | 5 | 95676039 | 95818985 | 496.0642 |

**pregnancy glycemic traits**
| **Pvalue** | **TopSNP** | **TopSNP-pvalue** |
| --- | --- | --- |
| 1.00E-06 | rs35261542 | 3.58E-12 |
| 1.00E-06 | rs10882085 | 1.74E-10 |
| 1.00E-06 | rs1573051 | 8.85E-11 |
| 1.00E-06 | rs3781637 | 2.04E-31 |
| 1.00E-06 | rs9348441 | 1.39E-30 |
| 1.00E-06 | rs11981584 | 2.82E-10 |
| 1.00E-06 | rs11981584 | 2.82E-10 |
| 2.00E-06 | rs10882096 | 1.30E-10 |
| 4.00E-06 | rs11716713 | 2.72E-06 |
| 6.00E-06 | rs76745696 | 5.33E-05 |
| 7.00E-06 | rs16951275 | 2.16E-06 |
| 9.00E-06 | rs147206241 | 1.79E-05 |
| 1.00E-05 | rs12439210 | 1.13E-06 |
| 1.00E-05 | rs73902879 | 2.35E-05 |
| 1.00E-06 | rs3781637 | 2.50E-18 |
| 1.00E-06 | rs35261542 | 1.94E-16 |
| 1.00E-06 | rs4731417 | 1.75E-07 |
| 1.00E-06 | rs4731417 | 1.75E-07 |
| 1.00E-06 | rs79257100 | 2.64E-11 |
| 1.00E-06 | rs3781637 | 1.08E-19 |
| 1.00E-06 | rs11716713 | 9.43E-10 |
| 1.00E-06 | rs35261542 | 2.36E-20 |
| 1.00E-06 | rs11981584 | 1.33E-06 |
| 1.00E-06 | rs11981584 | 1.33E-06 |
| 2.00E-06 | rs1573051 | 3.62E-10 |
| 2.00E-06 | rs74619378 | 1.31E-05 |
| 5.00E-06 | rs1044153 | 4.41E-10 |
| 9.00E-06 | rs2034345 | 1.29E-05 |
| 1.00E-06 | rs58433451 | 1.41E-12 |
| 1.00E-06 | rs58433451 | 1.41E-12 |
| 1.00E-06 | rs10734253 | 7.43E-09 |
| 1.00E-06 | rs74850326 | 3.40E-07 |
| 1.00E-06 | rs35261542 | 2.74E-18 |
| 2.00E-06 | rs58433451 | 1.41E-12 |
| 8.00E-06 | rs10734253 | 7.43E-09 |
| 1.00E-06 | rs3781637 | 8.37E-16 |
| 1.00E-06 | rs6048206 | 2.20E-11 |
| 1.00E-06 | rs6048206 | 2.20E-11 |
| 1.00E-06 | rs6048206 | 2.20E-11 |
| 1.00E-06 | rs12612019 | 1.09E-08 |
| 1.00E-06 | rs10213823 | 3.52E-11 |
| 1.00E-06 | rs9348441 | 1.54E-09 |
| 1.00E-06 | rs3217944 | 3.47E-13 |
| 1.00E-06 | rs3217944 | 3.47E-13 |
| 1.00E-06 | rs4731417 | 1.41E-06 |
| 1.00E-06 | rs4731417 | 1.41E-06 |
| 1.00E-06 | rs3217944 | 3.47E-13 |
| 1.00E-06 | rs3217944 | 3.47E-13 |
| 1.00E-06 | rs3217944 | 3.47E-13 |
| 1.00E-06 | rs75885458 | 4.95E-09 |
| 2.00E-06 | rs75070331 | 1.95E-08 |
| 4.00E-06 | rs12612019 | 1.09E-08 |
| 5.00E-06 | rs75353205 | 3.84E-06 |
| 7.00E-06 | rs3020430 | 1.29E-08 |
| 9.00E-06 | rs1042658 | 1.18E-06 |
| 1.00E-06 | rs10882085 | 2.43E-10 |
| 1.00E-06 | rs1573051 | 1.54E-10 |
| 1.00E-06 | rs3781637 | 5.27E-27 |
| 1.00E-06 | rs74619378 | 3.12E-06 |
| 1.00E-06 | rs11716713 | 5.69E-07 |
| 1.00E-06 | rs35261542 | 7.51E-25 |
| 1.00E-06 | rs11981584 | 5.36E-09 |
| 1.00E-06 | rs11981584 | 5.36E-09 |
| 3.00E-06 | rs10882096 | 2.26E-10 |
| 5.00E-06 | rs12439210 | 1.43E-06 |
| 1.00E-05 | rs1946701 | 0.000215841 |
| 1.00E-06 | rs58433451 | 6.54E-09 |
| 1.00E-06 | rs3781637 | 4.72E-18 |
| 1.00E-06 | rs9348441 | 4.46E-25 |
| 1.00E-06 | rs11981584 | 2.10E-07 |
| 1.00E-06 | rs11981584 | 2.10E-07 |
| 2.00E-06 | rs58433451 | 6.54E-09 |
| 8.00E-06 | rs59139497 | 6.07E-07 |

**Table S4.**
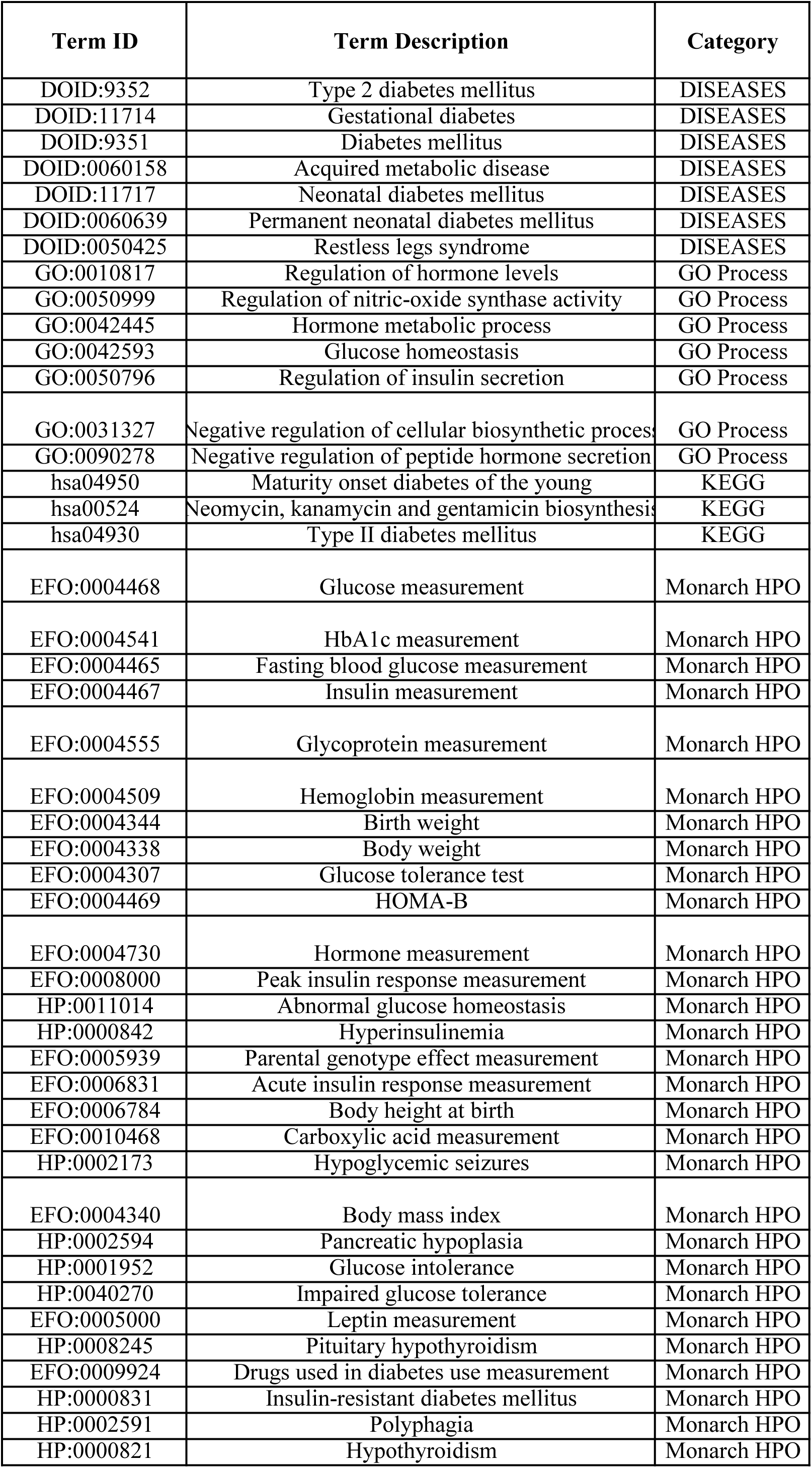
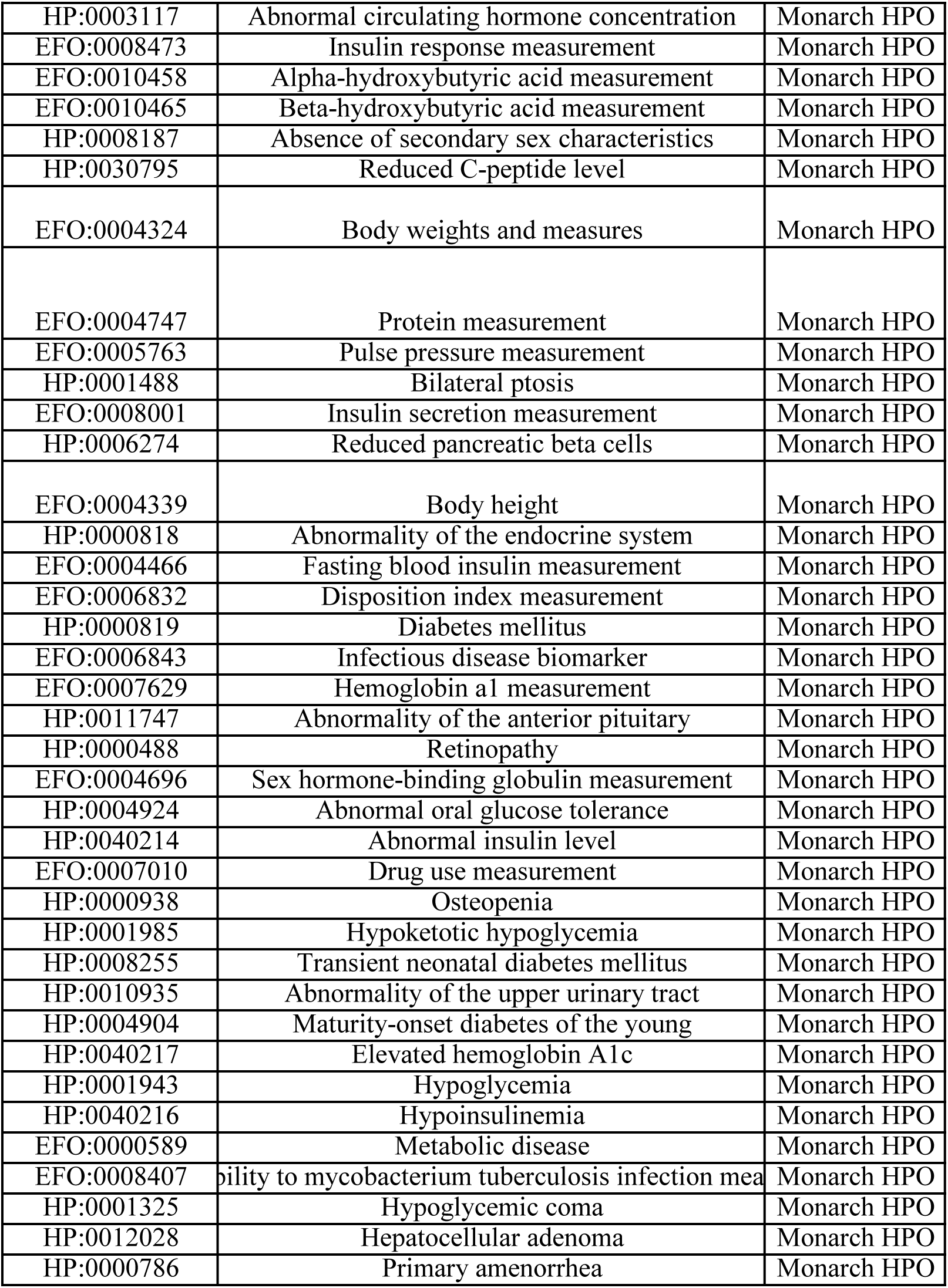
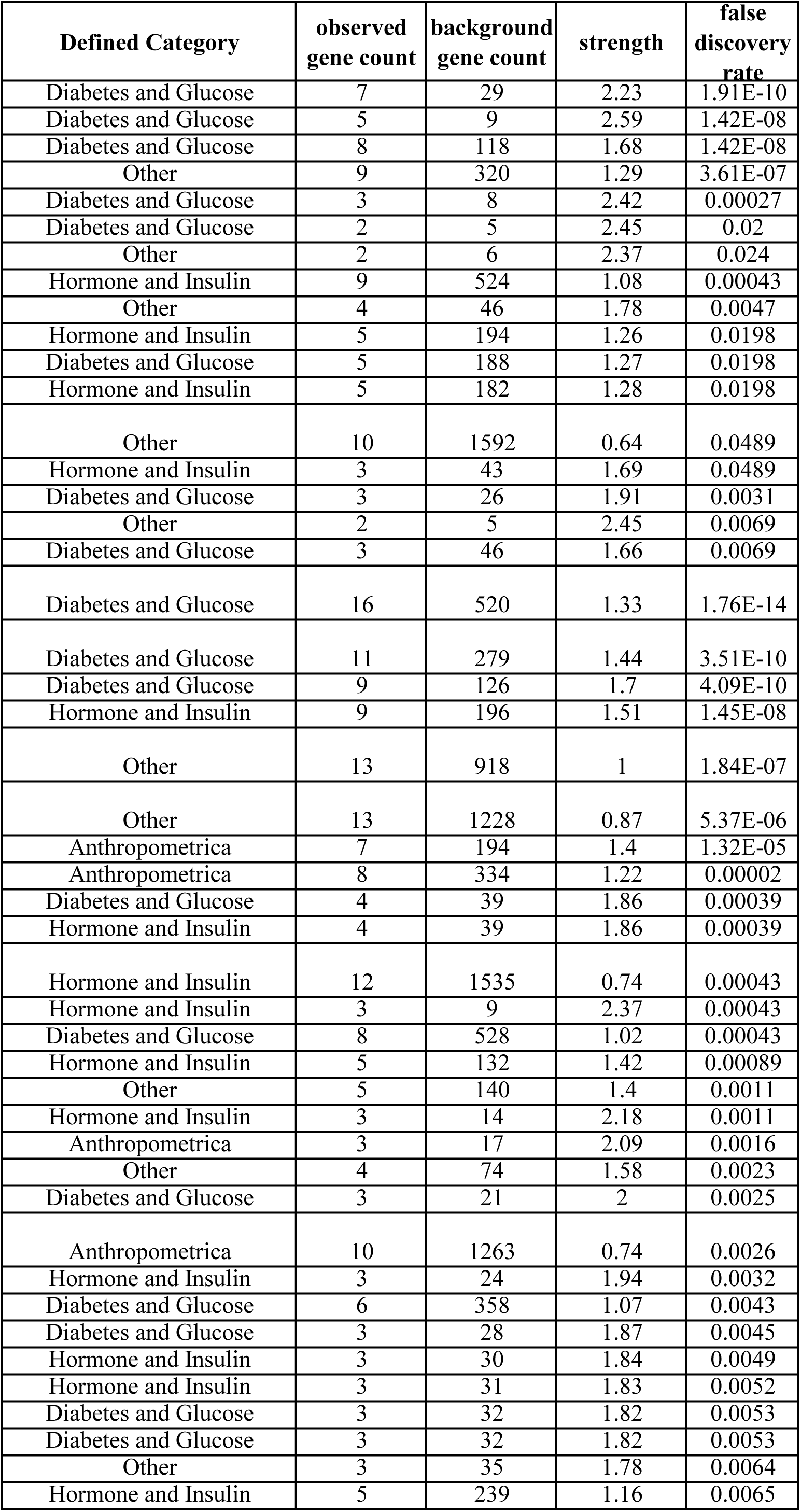
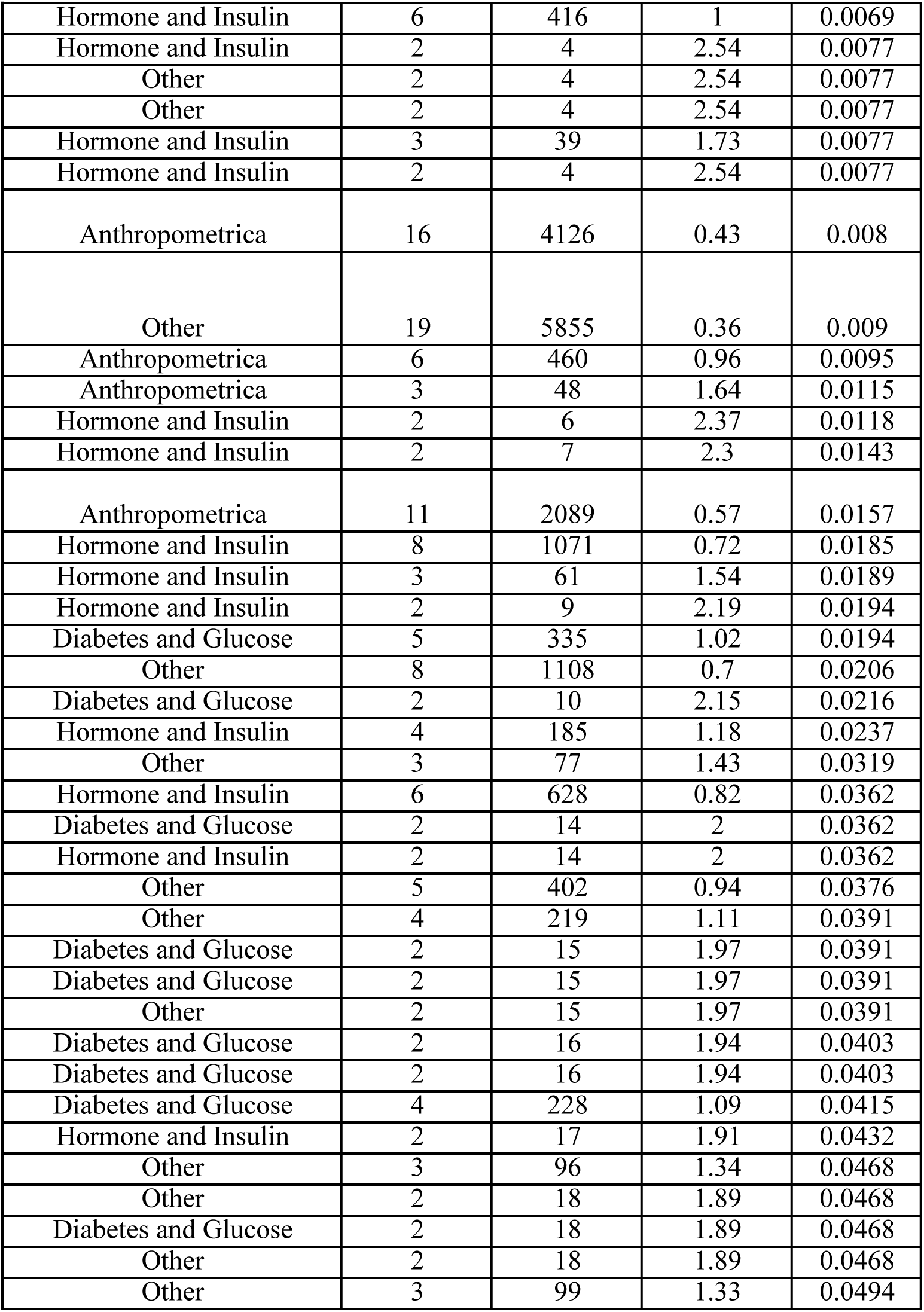
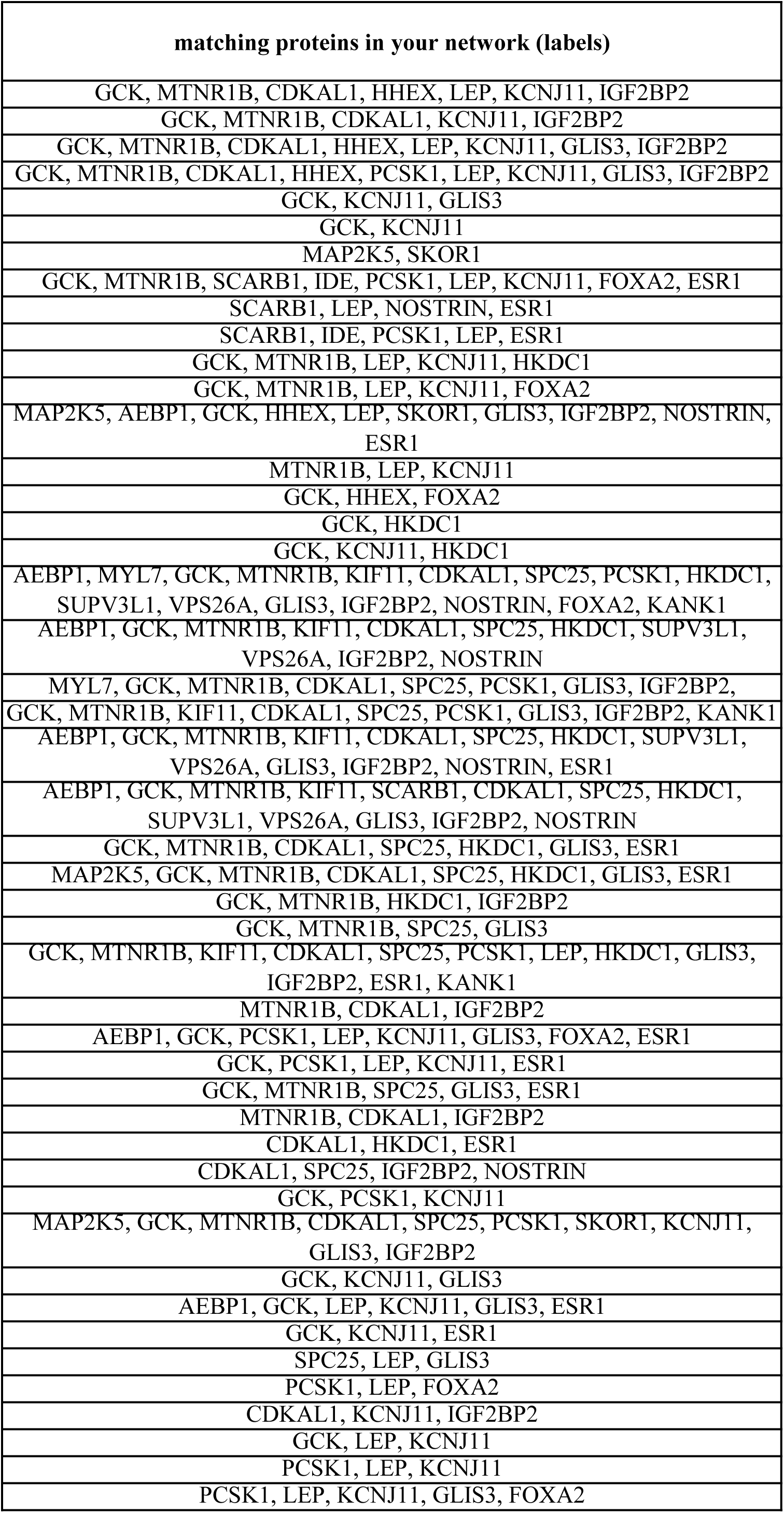
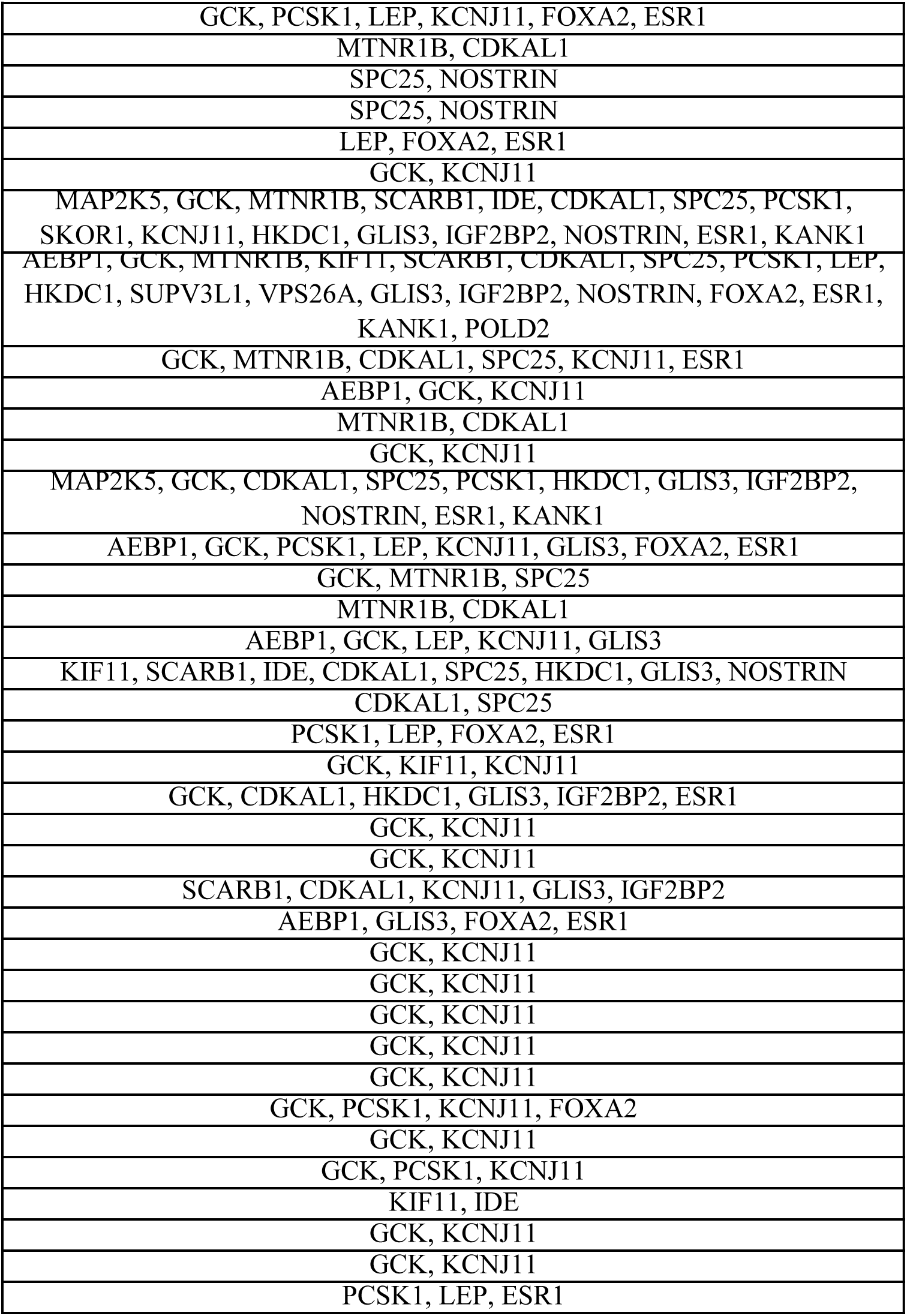
The STRING functional enrichment analysis results.

## References

1 McIntyre, H. D. et al. Gestational diabetes mellitus. Nat Rev Dis Primers 5, 47, doi:10.1038/s41572-019-0098-8 (2019).

2 Gao, C., Sun, X., Lu, L., Liu, F. & Yuan, J. Prevalence of gestational diabetes mellitus in mainland China: A systematic review and meta-analysis. J Diabetes Investig 10, 154–162, doi:10.1111/jdi.12854 (2019).

3 Buchanan, T. A., Xiang, A. H. & Page, K. A. Gestational diabetes mellitus: risks and management during and after pregnancy. Nat Rev Endocrinol 8, 639–649, doi:10.1038/nrendo.2012.96 (2012).

4 Weissgerber, T. L. & Mudd, L. M. Preeclampsia and diabetes. Current diabetes reports 15, 9, doi:10.1007/s11892-015-0579-4 (2015).

5 Gabbay-Benziv, R., Reece, E. A., Wang, F. & Yang, P. Birth defects in pregestational diabetes: Defect range, glycemic threshold and pathogenesis. World J Diabetes 6, 481–488, doi:10.4239/wjd.v6.i3.481 (2015).

6 in IDF Diabetes Atlas, 9th edn. (International Diabetes Federation © International Diabetes Federation, 2019., 2019).

7 Metzger, B. E. et al. Hyperglycemia and adverse pregnancy outcomes. The New England journal of medicine 358, 1991–2002, doi:10.1056/NEJMoa0707943 (2008).

8 Morisset, A. S. et al. Prevention of gestational diabetes mellitus: a review of studies on weight management. Diabetes Metab Res Rev 26, 17–25, doi:10.1002/dmrr.1053 (2010).

9 Reece, E. A. The fetal and maternal consequences of gestational diabetes mellitus. The journal of maternal-fetal & neonatal medicine : the official journal of the European Association of Perinatal Medicine, the Federation of Asia and Oceania Perinatal Societies, the International Society of Perinatal Obstet 23, 199–203, doi:10.3109/14767050903550659 (2010).

10 Petry, C. J. Gestational diabetes: risk factors and recent advances in its genetics and treatment. Br J Nutr 104, 775–787, doi:10.1017/s0007114510001741 (2010).

11 Ozaki, K. et al. Functional SNPs in the lymphotoxin-alpha gene that are associated with susceptibility to myocardial infarction. Nature genetics 32, 650–654, doi:10.1038/ng1047 (2002).

12 Kwak, S. H. et al. A genome-wide association study of gestational diabetes mellitus in Korean women. Diabetes 61, 531–541, doi:10.2337/db11-1034 (2012).

13 Wu, N. N. et al. A genome-wide association study of gestational diabetes mellitus in Chinese women. The journal of maternal-fetal & neonatal medicine : the official journal of the European Association of Perinatal Medicine, the Federation of Asia and Oceania Perinatal Societies, the International Society of Perinatal Obstet 34, 1557–1564, doi:10.1080/14767058.2019.1640205 (2021).

14 Backman, J. D. et al. Exome sequencing and analysis of 454,787 UK Biobank participants. Nature 599, 628–634, doi:10.1038/s41586-021-04103-z (2021).

15 Jiang, L., Zheng, Z., Fang, H. & Yang, J. A generalized linear mixed model association tool for biobank-scale data. Nature genetics 53, 1616–1621, doi:10.1038/s41588-021-00954-4 (2021).

16 Pervjakova, N. et al. Multi-ancestry genome-wide association study of gestational diabetes mellitus highlights genetic links with type 2 diabetes. Hum Mol Genet 31, 3377–3391, doi:10.1093/hmg/ddac050 (2022).

17 Changalidis, A. I. et al. Aggregation of Genome-Wide Association Data from FinnGen and UK Biobank Replicates Multiple Risk Loci for Pregnancy Complications. Genes (Basel) 13, doi:10.3390/genes13122255 (2022).

18 Benn, P. et al. Position statement from the Aneuploidy Screening Committee on behalf of the Board of the International Society for Prenatal Diagnosis. Prenat Diagn 33, 622–629, doi:10.1002/pd.4139 (2013).

19 Liu, S. et al. Genomic Analyses from Non-invasive Prenatal Testing Reveal Genetic Associations, Patterns of Viral Infections, and Chinese Population History. Cell 175, 347–359.e314, doi:10.1016/j.cell.2018.08.016 (2018).

20 Congyue Zhang, Y. W., Weijie Sun, Huixia Yang. The Area under the Curve (AUC) of Oral Glucose Tolerance Test (OGTT) Could Be a Measure Method of Hyperglycemia in All Pregnant Women. Open Journal of Obstetrics and Gynecology 9, 186–195, doi:10.4236/ojog.2019.92019. (2019).

21 Pascoe, L. et al. Common variants of the novel type 2 diabetes genes CDKAL1 and HHEX/IDE are associated with decreased pancreatic beta-cell function. Diabetes 56, 3101–3104, doi:10.2337/db07-0634 (2007).

22 Steinthorsdottir, V. et al. A variant in CDKAL1 influences insulin response and risk of type 2 diabetes. Nature genetics 39, 770–775, doi:10.1038/ng2043 (2007).

23 Rubio-Sastre, P., Scheer, F. A., Gómez-Abellán, P., Madrid, J. A. & Garaulet, M. Acute melatonin administration in humans impairs glucose tolerance in both the morning and evening. Sleep 37, 1715–1719, doi:10.5665/sleep.4088 (2014).

24 Mao, X. et al. The deep population history of northern East Asia from the Late Pleistocene to the Holocene. Cell 184, 3256–3266.e3213, doi:10.1016/j.cell.2021.04.040 (2021).

25 Ling, Y. et al. A common polymorphism rs3781637 in MTNR1B is associated with type 2 diabetes and lipids levels in Han Chinese individuals. Cardiovasc Diabetol 10, 27, doi:10.1186/1475-2840-10-27 (2011).

26 Lai, I. C. et al. Analysis of genetic variations in the human melatonin receptor (MTNR1A, MTNR1B) genes and antipsychotics-induced tardive dyskinesia in schizophrenia. World J Biol Psychiatry 12, 143–148, doi:10.3109/15622975.2010.496870 (2011).

27 Dietrich, K. et al. Association and evolutionary studies of the melatonin receptor 1B gene (MTNR1B) in the self-contained population of Sorbs from Germany. Diabet Med 28, 1373–1380, doi:10.1111/j.1464-5491.2011.03374.x (2011).

28 Maugars, G., Nourizadeh-Lillabadi, R. & Weltzien, F. A. New Insights Into the Evolutionary History of Melatonin Receptors in Vertebrates, With Particular Focus on Teleosts. Frontiers in endocrinology 11, 538196, doi:10.3389/fendo.2020.538196 (2020).

29 Song, J. F. et al. Evaluation of the effect of MTNR1B rs10830963 gene variant on the therapeutic efficacy of nateglinide in treating type 2 diabetes among Chinese Han patients. BMC Med Genomics 14, 156, doi:10.1186/s12920-021-01004-y (2021).

30 Li, Z. et al. CMDB: the comprehensive population genome variation database of China. Nucleic acids research 51, D890–d895, doi:10.1093/nar/gkac638 (2023).

31 Mahajan, A. et al. Multi-ancestry genetic study of type 2 diabetes highlights the power of diverse populations for discovery and translation. Nature genetics 54, 560–572, doi:10.1038/s41588-022-01058-3 (2022).

32 Hayes, M. G. et al. Identification of HKDC1 and BACE2 as genes influencing glycemic traits during pregnancy through genome-wide association studies. Diabetes 62, 3282–3291, doi:10.2337/db12-1692 (2013).

33 Kim, Y. J. et al. The contribution of common and rare genetic variants to variation in metabolic traits in 288,137 East Asians. Nature communications 13, 6642, doi:10.1038/s41467-022-34163-2 (2022).

34 Lee, C. J. et al. Phenome-wide analysis of Taiwan Biobank reveals novel glycemia-related loci and genetic risks for diabetes. Commun Biol 5, 1175, doi:10.1038/s42003-022-04168-0 (2022).

35 Richardson, T. G. et al. Characterising metabolomic signatures of lipid-modifying therapies through drug target mendelian randomisation. PLoS biology 20, e3001547, doi:10.1371/journal.pbio.3001547 (2022).

36 Morris, A. P. et al. Large-scale association analysis provides insights into the genetic architecture and pathophysiology of type 2 diabetes. Nature genetics 44, 981–990, doi:10.1038/ng.2383 (2012).

37 Kichaev, G. et al. Leveraging Polygenic Functional Enrichment to Improve GWAS Power. Am J Hum Genet 104, 65–75, doi:10.1016/j.ajhg.2018.11.008 (2019).

38 Suzuki, K. et al. Identification of 28 new susceptibility loci for type 2 diabetes in the Japanese population. Nature genetics 51, 379–386, doi:10.1038/s41588-018-0332-4 (2019).

39 Zhu, Z. et al. Shared genetic and experimental links between obesity-related traits and asthma subtypes in UK Biobank. The Journal of allergy and clinical immunology 145, 537–549, doi:10.1016/j.jaci.2019.09.035 (2020).

40 Sun, J. W., Collins, J. M., Ling, D. & Wang, D. Highly Variable Expression of ESR1 Splice Variants in Human Liver: Implication in the Liver Gene Expression Regulation and Inter-Person Variability in Drug Metabolism and Liver Related Diseases. J Mol Genet Med 13 (2019).

41 Johns, E. C., Denison, F. C., Norman, J. E. & Reynolds, R. M. Gestational Diabetes Mellitus: Mechanisms, Treatment, and Complications. Trends Endocrinol Metab 29, 743–754, doi:10.1016/j.tem.2018.09.004 (2018).

42 Li, C. et al. Association of Estrogen Receptor α Gene Polymorphism and its Expression with Gestational Diabetes Mellitus. Gynecol Obstet Invest 85, 26–33, doi:10.1159/000502378 (2020).

43 Yang, S., Shi, F. T., Leung, P. C., Huang, H. F. & Fan, J. Low Thyroid Hormone in Early Pregnancy Is Associated With an Increased Risk of Gestational Diabetes Mellitus. J Clin Endocrinol Metab 101, 4237–4243, doi:10.1210/jc.2016-1506 (2016).

44 Cooray, S. D. et al. Development, validation and clinical utility of a risk prediction model for adverse pregnancy outcomes in women with gestational diabetes: The PeRSonal GDM model. EClinicalMedicine 52, 101637, doi:10.1016/j.eclinm.2022.101637 (2022).

45 Kawai, V. K. et al. A genetic risk score that includes common type 2 diabetes risk variants is associated with gestational diabetes. Clinical endocrinology 87, 149–155, doi:10.1111/cen.13356 (2017).

46 Wu, Q. et al. An early prediction model for gestational diabetes mellitus based on genetic variants and clinical characteristics in China. Diabetol Metab Syndr 14, 15, doi:10.1186/s13098-022-00788-y (2022).

47 Sakaguchi, K. et al. Glucose area under the curve during oral glucose tolerance test as an index of glucose intolerance. Diabetology international 7, 53–58, doi:10.1007/s13340-015-0212-4 (2016).

48 Youden, W. J. Index for rating diagnostic tests. Cancer 3, 32–35, doi:10.1002/1097-0142(1950)3:1<32::aid-cncr2820030106>3.0.co;2-3 (1950).

49 Kc, C., Y, L. & Cheng, J.-T. The Areas Under Curves (AUC) used in diabetes research: Update view. Integrative Obesity and Diabetes 4, doi:10.15761/IOD.1000212 (2018).

50 Chang, C. C., et al. Second-generation PLINK: rising to the challenge of larger and richer datasets. Gigascience 4, 7, doi:10.1186/s13742-015-0047-8 (2015).

51 Yang, J. et al. Conditional and joint multiple-SNP analysis of GWAS summary statistics identifies additional variants influencing complex traits. Nature genetics 44, 369–375, s361-363, doi:10.1038/ng.2213 (2012).

52 Yang, J., Lee, S. H., Goddard, M. E. & Visscher, P. M. GCTA: a tool for genome-wide complex trait analysis. Am J Hum Genet 88, 76–82, doi:10.1016/j.ajhg.2010.11.011 (2011).

53 Mishra, A. & Macgregor, S. VEGAS2: Software for More Flexible Gene-Based Testing. Twin research and human genetics : the official journal of the International Society for Twin Studies 18, 86–91, doi:10.1017/thg.2014.79 (2015).

54 Snel, B., Lehmann, G., Bork, P. & Huynen, M. A. STRING: a web-server to retrieve and display the repeatedly occurring neighbourhood of a gene. Nucleic acids research 28, 3442–3444, doi:10.1093/nar/28.18.3442 (2000).

55 Guo, X. et al. CNSA: a data repository for archiving omics data. Database : the journal of biological databases and curation 2020, doi:10.1093/database/baaa055 (2020).

56 Chen, F. Z. et al. CNGBdb: China National GeneBank DataBase. Yi chuan = Hereditas 42, 799–809, doi:10.16288/j.yczz.20-080 (2020).

